# Navigating the Cholera Elimination Roadmap in Zambia - a Scoping Review (2013-2023)

**DOI:** 10.1101/2024.08.05.24311486

**Authors:** Nyuma Mbewe, John Tembo, Mpanga Kasonde, Kelvin Mwangilwa, Paul Zulu, Joseph Sereki, William Ngosa, Kennedy Lishipmi, Lloyd Mulenga, Roma Chilengi, Nathan Kapata, Martin P. Grobusch

**Affiliations:** National Cholera Elimination Taskforce, Zambia National Public Health Institute, Lusaka, Zambia; HERPEZ Zambia – Institute for Infectious Disease Research, Lusaka, Zambia; Regional Cholera Support Coordinator, International Federation of Red Cross Society, Lusaka, Zambia; Ministry of Health Headquarters, Ndeke House, Lusaka, Zambia; Center of Tropical Medicine and Travel Medicine, Department of Infectious Diseases, Amsterdam University Medical Centres, location AMC, Amsterdam Infection & Immunity, Amsterdam Public Health, University of Amsterdam, Amsterdam, The Netherlands

**Keywords:** Cholera, GTFCC, Cholera control, elimination, Climate Change, Zambia, sub-Saharan Africa

## Abstract

**Background:** Cholera outbreaks are increasing in frequency and severity, particularly in Sub-Saharan Africa. Zambia, committed to ending cholera by 2025, is coming off its most significant outbreak in 2024. This review examines the perceived regression in elimination efforts by addressing two questions: (1) what is known about cholera in Zambia; and (2) what are the main suggested mechanisms and strategies to further elimination efforts in the region?

**Methodology/Principal Findings:** A scoping literature search was conducted in PUBMED to identify relevant studies published between January 2013 and June 2024 using the search terms ‘cholera’ and ‘Zambia’. We identified 45 relevant publications. With the increasing influence of climate change, population growth, and rural-urban migration, further increases in outbreak frequency and magnitude are expected. Major risk factors for recurrent outbreaks include poor access to water, sanitation, and hygiene services in urban unplanned settlements and rural fishing villages. Interventions are best planned at a decentralized, community-centric approach to prevent elimination and reintroduction at the district level. Pre-emptive vaccination campaigns before the rainy season and climate-resilient WASH infrastructure are also recommended.

**Conclusions/Significance:** The goal to eliminate cholera by 2025 was unrealistic as evidence points to the disease becoming endemic. Our findings confirm the need to align health and WASH investments with the Global Roadmap to Cholera Elimination by 2030 through a climate-focused lens. Recommendations for cholera elimination, including improved access to safe drinking water and sanitation, remain elusive in many low-income settings like Zambia. Patient-level information on survival and transmissibility is lacking. New research tailored to country-level solutions is urgently required. Insights from this review will be integrated into the next iteration of the National Cholera Control Plan and could be applicable to other countries with similar settings.

**Article Summary:** Despite known evidence of the risks from insufficient safe water supplies, sanitation and hygiene (WASH), the protective effects of oral cholera vaccines, and a Roadmap from the Global Task Force on Cholera Control, there is a continuous increase in cholera outbreaks on the continent. Now endemic in many parts of Zambia, it is postulated that the true burden of cholera in the country is underreported due to inadequate completeness of data, particularly during outbreaks. With an increasing frequency related to climatic conditions and unplanned urbanization, it will be important to adopt a decentralised approach to cholera control in Zambia. There is a continued need to advocate strongly for multisectoral interventions aligning health and WASH investments. The findings expose gaps in the local literature, such as how to improve climate-resilient WASH infrastructure, strategies to boost vaccine availability, and also the host and environmental factors that may be protective at personal and household levels from being asymptomatic or dying of cholera. This work provides evidence-based recommendations for the next iteration of the National Cholera Control Plan for Zambia and for neighbouring countries that may be in the process of developing their own plans.

## Introduction

Cholera outbreaks are increasing in frequency and severity across the world, particularly in sub-Saharan Africa. This is despite efforts by the Global Task Force on Cholera Control (GTFCC) to achieve cholera elimination in at least 20 countries by 2030 [1]. Zambia, with its Republican President serving as the Global Champion for Cholera Control, had set out to lead the elimination efforts by 2025 ahead of the global targets [2]. However, the country is now coming off experiencing its most significant outbreak to date with 23,381 cumulative cases, and 740 fatalities of which 304 were facility deaths representing a case fatality of 1.8% (Accessed on 31^st^ July 2024 [3]. We reported elsewhere a survival analysis of a cohort of patients admitted to treatment centres in Lusaka and found that lack of prior vaccination and the presence of comorbidities were statistically significant contributors to in-patient mortality [4].

The GTFCC Roadmap to Cholera Elimination by 2030 focuses on investment in Water Sanitation and Hygiene (WASH), early case investigation, and the systematic use of Oral Cholera Vaccines (OCV) as part of cholera elimination strategies as a bridge towards longer-term investments in Water Sanitation and Hygiene (WASH) [1]. The country is currently conducting a mid-term revision of the National Cholera Control Plan (NCP). It was thus necessary to undertake this work in order to understand what constitutes published knowledge on cholera in Zambia and also to learn from lessons and evidence-based practices that could contribute to reduced cholera mortality and overall number of cases in outbreaks by 2030.

Several other countries earmarked for cholera elimination have documented progress and lessons learned. Haiti, for example, notes the need for case-area targeted interventions, given ongoing vulnerabilities and vaccine shortages [5]. In the Democratic Republic of Congo (DRC), a narrative review detailed the successes and challenges in the implementation of three iterations of their multisectoral cholera elimination plan (2008-2012, 2013-2017 and 2018-2021) to influence the implementation of their NCP 2023-2027 [6]. They noted that there has been largely no change since the pre-NCP period. Lastly, Uganda noted the use of a scorecard to track cholera elimination efforts at district and ward levels. They highlighted the risks of periods of elimination and then resurgence in some areas if ongoing elimination efforts such as improved WASH and OCV campaigns were not sustained [7]. Global efforts to improve vaccine availability and rapid diagnostic kits must be matched by domestic adaptation of GTFCC guidelines to ensure better response efforts during outbreaks and a speedier transition from control to elimination of cholera in endemic countries.

To better adapt cholera control and elimination strategies in Zambia, we summarized the current evidence on the cholera situation in the country. With a One-Health approach of interaction between human, animal and environmental factors, this scoping review was undertaken to summarise existing evidence on cholera epidemiology and elimination in Zambia. By examining the perceived regression in cholera elimination efforts, we sought to document the evidence generated from the different pillars to facilitate a comprehensive multisectoral response strategy. We addressed two main questions: (1) what is known about cholera in Zambia; (2) what are the main suggested mechanisms and strategies to further cholera control efforts in the region?

## Methods

A scoping literature search was conducted to identify relevant studies. Our goal was to map the existing literature, present evidence-based strategies in the different thematic areas of prevention/control, and present hypotheses on the best strategy to accelerate progress towards cholera control and eventual elimination in Zambia. We also sought to identify gaps in the research data that could be important for prioritizing intervention areas which may otherwise be overlooked.

In preparation for this narrative review, articles were identified in PUBMED using the search terms ‘cholera’ and ‘Zambia’ for articles published between 1^st^ January 2013 and 30^th^ May 2024; filtered to English only. We used the same search terms for Embase and Google Scholar. Reference lists of selected papers and reviews were also screened for relevant papers, as were local publications and preprints within the period under review. We excluded meeting reports, conference proceedings, daily situation reports, study protocols and articles related to cholera but not specifically about Zambia. The search was conducted, and all papers were screened between December 2023 and May 2024.

## Results

### Study Identification and Selection

A total of 45 records were identified that investigated cholera and Zambia from 2013 – 2024 from PUBMED, including one previous article exploring the epidemiology of cholera in Zambia from 2000-2010 [8]. Embase was inaccessible due to institutional restrictions, whilst Google Scholar brought up 11,300 articles. Eight studies that did not mention Zambia in the main text were excluded; and six additional ones were identified from alternative sources such as in preprint, in local journals or otherwise not listed on PUBMED were included [9–13]. Full texts were available for all the studies and reported according to the PRISMA guidelines [14] (Figure 1). All identified citations were uploaded into a Mendeley database and data was extracted using a predesigned form. Key findings and study designs were then collated into thematic areas based on the GTFCC Global Road Map for Cholera Control [1]. The GTFCC describes three axes achievable across six different pillars for a comprehensive multisectoral control plan [1]. Figure 2 shows how the analysed publications were evaluated in light of the different pillars.

**Figure 1:**
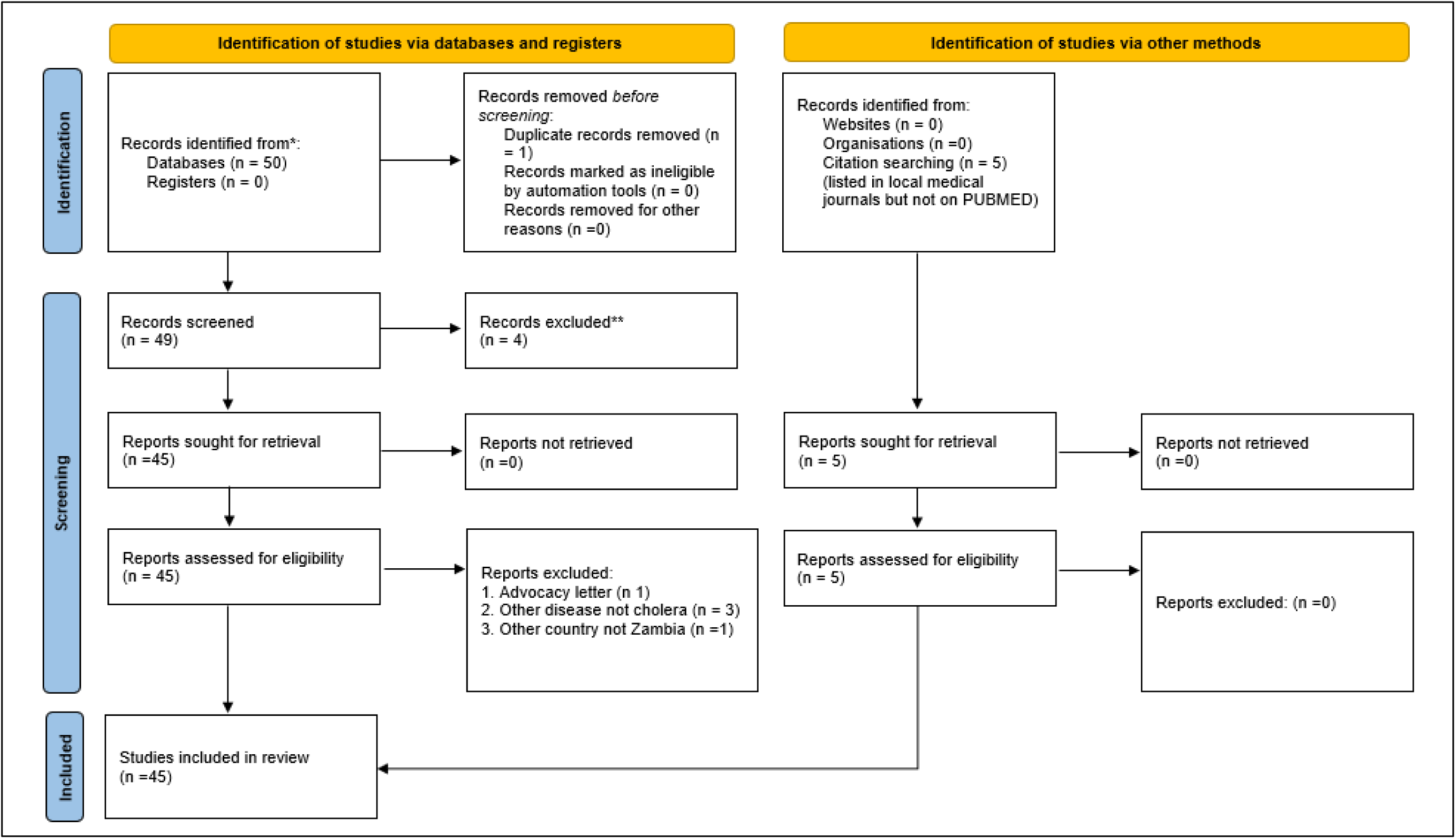
Study flow diagram showing the selection processes in line with the Prisma Guidelines.

**Figure 2:**
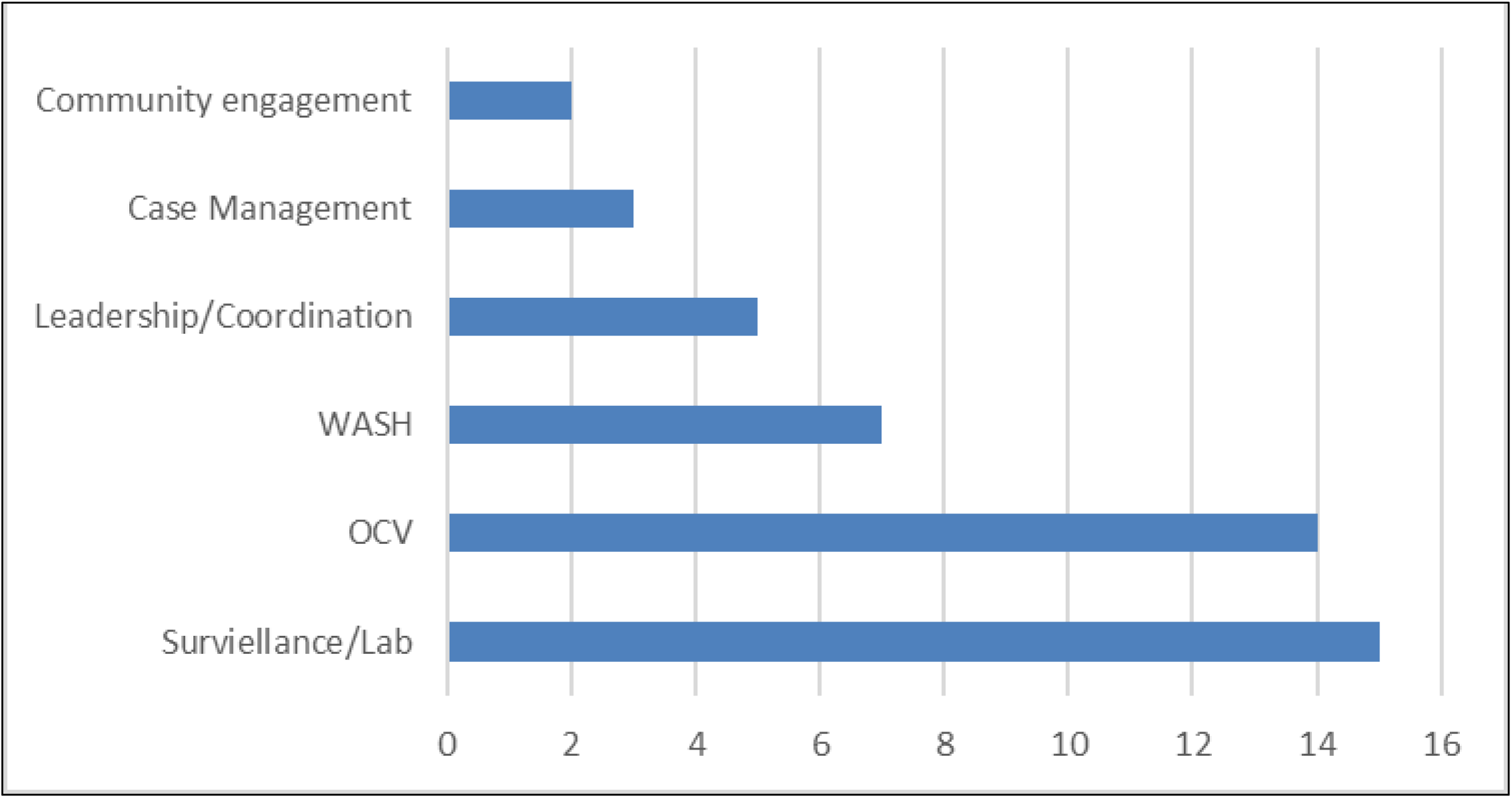
Frequency of Publications by Pillar.

### Cholera Epidemiology and Burden

Table 1 shows the overall epidemiology of cholera in Zambia. Over the years and consistently across the studies, over 80% of the cases were identified in peri-urban areas, particularly in Lusaka – the capital. Consistently, the definition of an outbreak, based on the national Integrated Disease Surveillance and Response (IDSR) guidelines, was the confirmation by stool culture of *V. cholerae* in at least one cholera suspect patient with three episodes of acute watery diarrhoea in a 24-hour period in each district [15]. Subsequent cholera suspect patients are included in the total case count per year based on fitting the clinical case definition with or without culture confirmation. All the referenced papers document cholera epidemiology in Zambia during outbreaks and show an increasing annual incidence [8,16,17]. Given the endemicity in certain parts of the country, it has been postulated that by only exploring the proportion of clinically suspected cholera cases and not all confirmed by culture, the true incidence of *V. cholerae* infections in the region and the true burden of disease may be higher, than reported especially outside outbreak seasons [18].

**Table 1:**
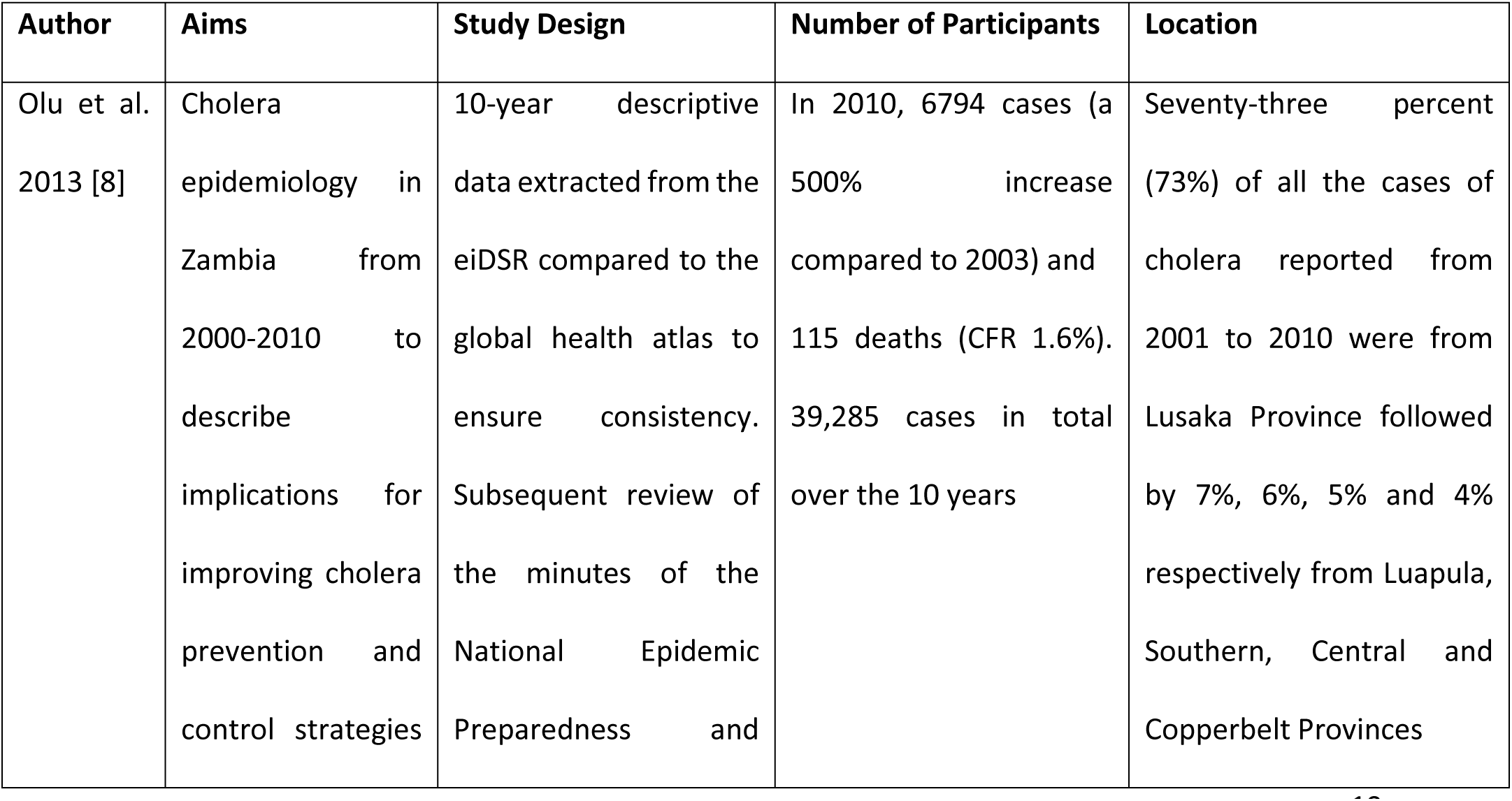
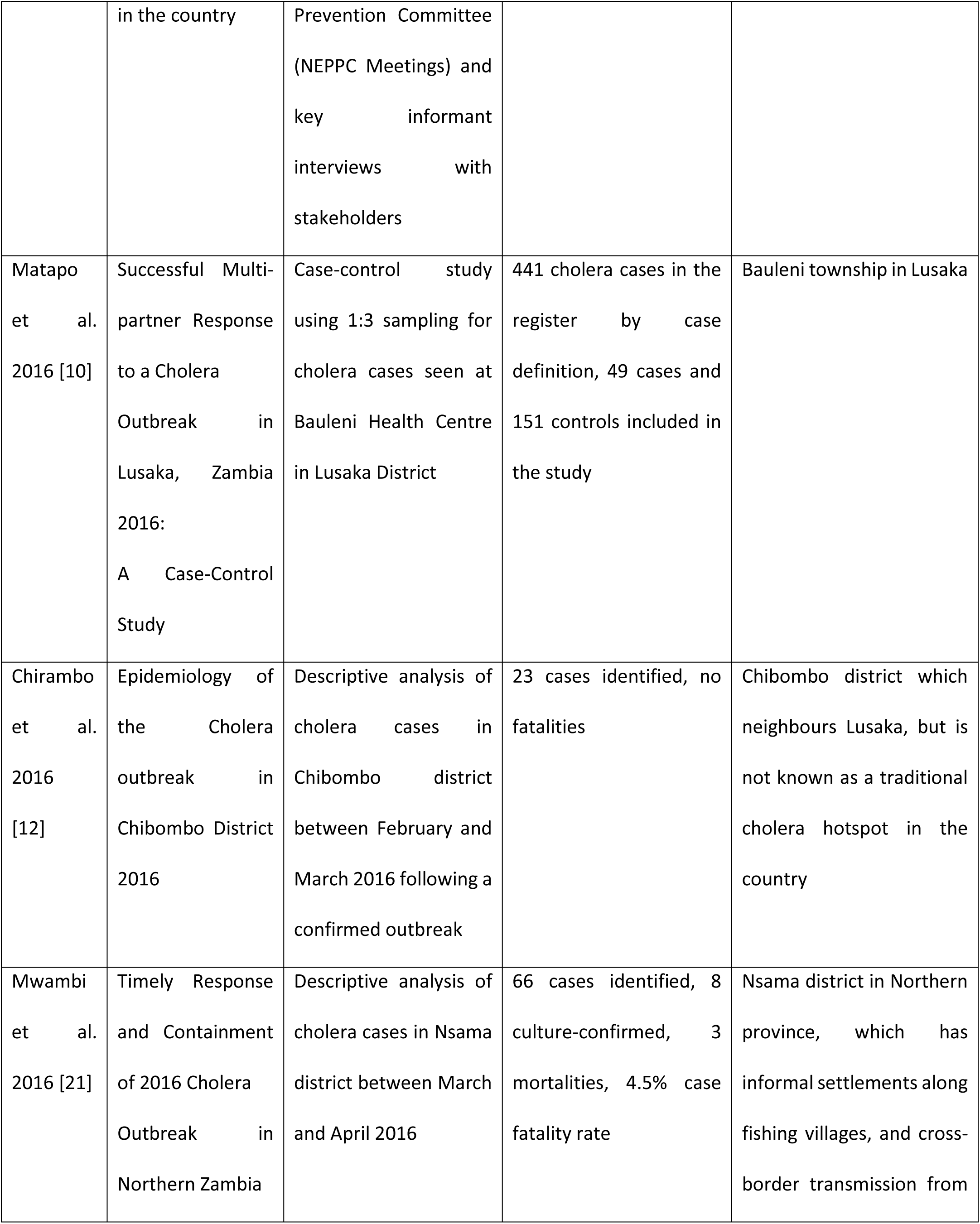
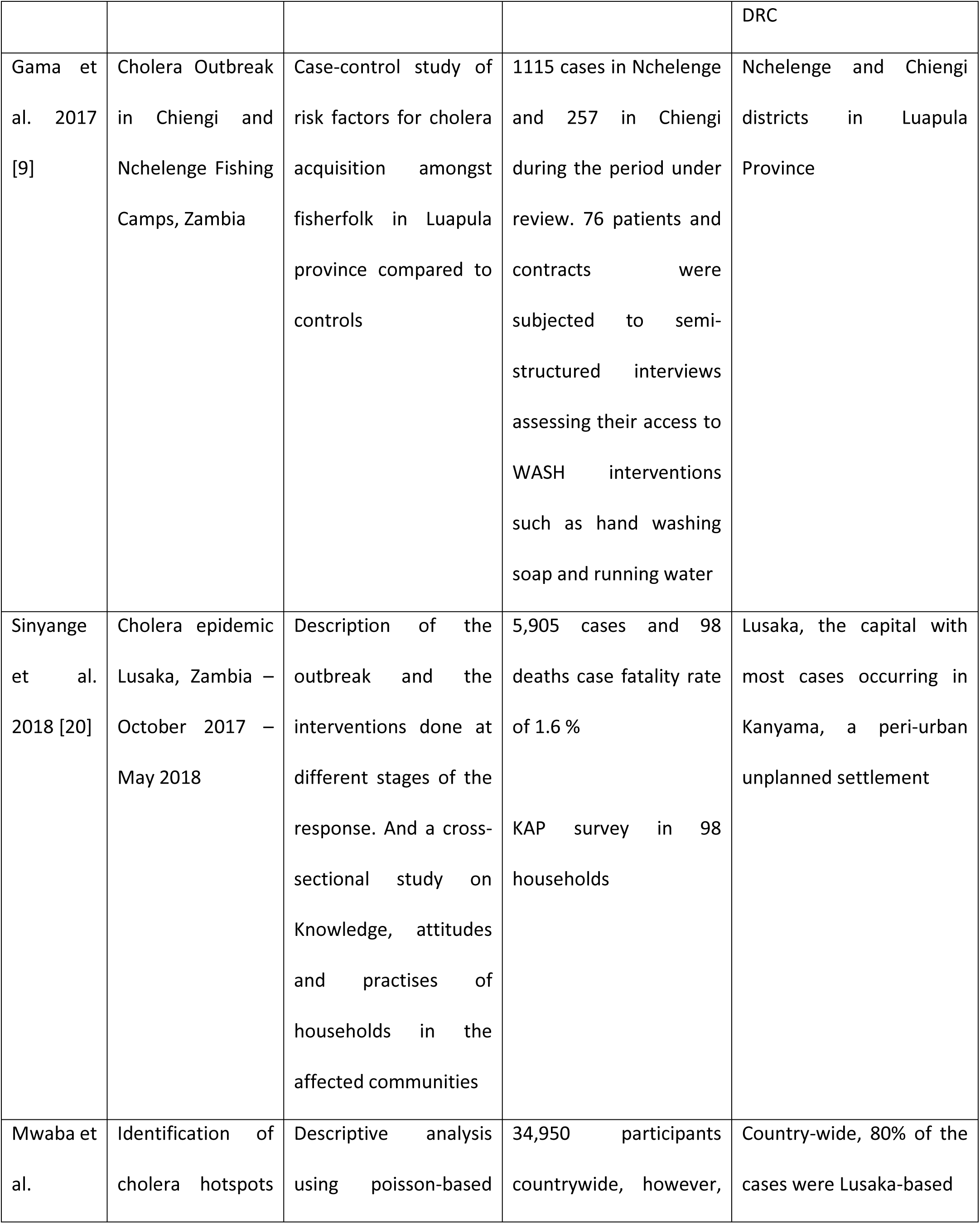
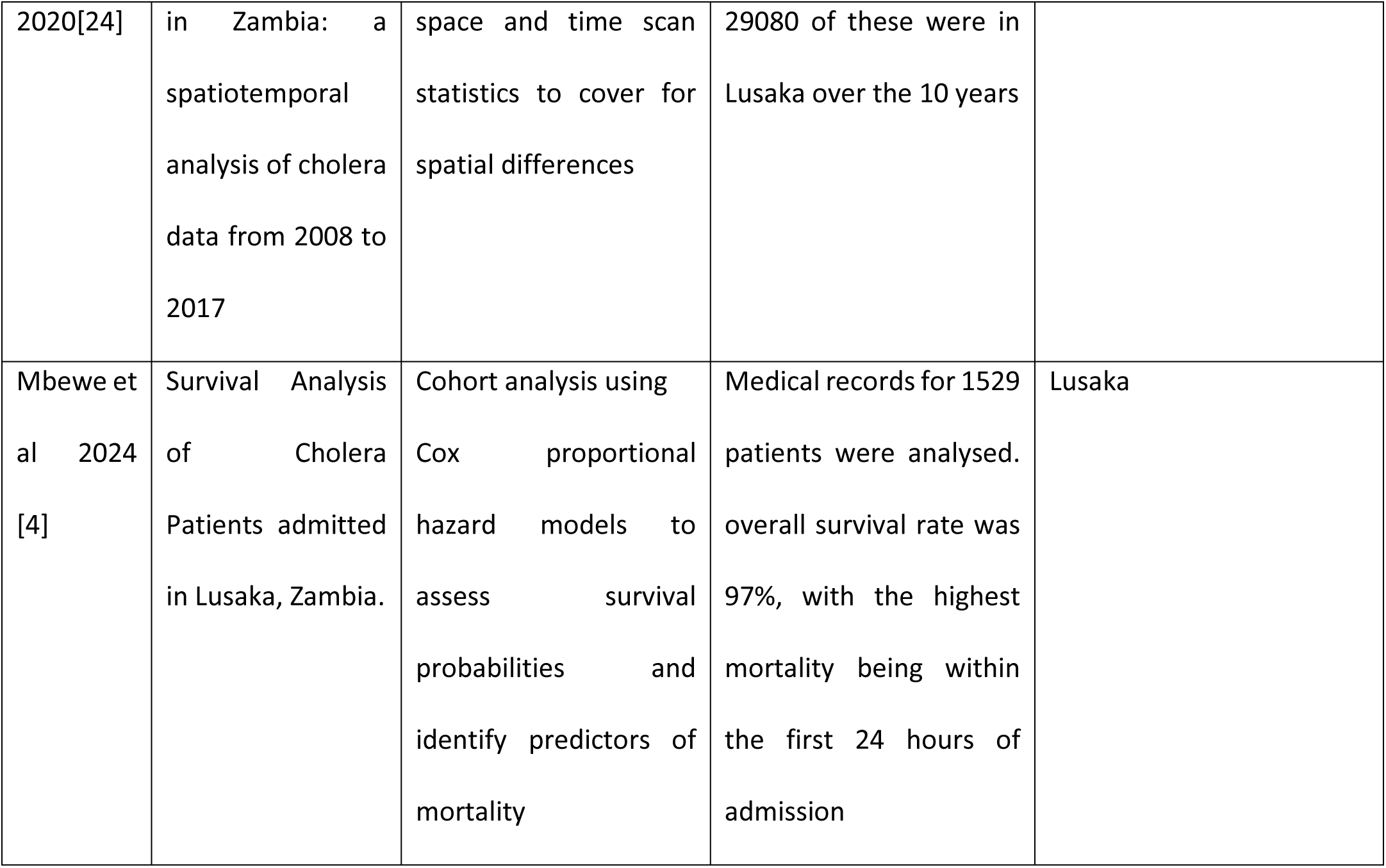
Cholera Epidemiology and Burden in Zambia (2013-2024)

| Author | Aims | Study Design | Number of Participants | Location |
| --- | --- | --- | --- | --- |
| Olu et al.<br>2013 [8] | Cholera epidemiology in Zambia from 2000-2010 to describe implications for improving cholera prevention and control strategies | 10-year descriptive data extracted from the eiDSR compared to the global health atlas to ensure consistency. Subsequent review of the minutes of the National Epidemic Preparedness and | In 2010, 6794 cases (a 500% increase compared to 2003) and 115 deaths (CFR 1.6%). 39,285 cases in total over the 10 years | Seventy-three percent (73%) of all the cases of cholera reported from 2001 to 2010 were from Lusaka Province followed by 7%, 6%, 5% and 4% respectively from Luapula, Southern, Central and Copperbelt Provinces |
|  | in the country | Prevention Committee (NEPPC Meetings) and key informant interviews with stakeholders |  |  |
| Matapo et al. 2016 [10] | Successful Multi-partner Response to a Cholera Outbreak in Lusaka, Zambia 2016: A Case-Control Study | Case-control study using 1:3 sampling for cholera cases seen at Bauleni Health Centre in Lusaka District | 441 cholera cases in the register by case definition, 49 cases and 151 controls included in the study | Bauleni township in Lusaka |
| Chirambo et al. 2016 [12] | Epidemiology of the Cholera outbreak in Chibombo District 2016 | Descriptive analysis of cholera cases in Chibombo district between February and March 2016 following a confirmed outbreak | 23 cases identified, no fatalities | Chibombo district which neighbours Lusaka, but is not known as a traditional cholera hotspot in the country |
| Mwambi et al. 2016 [21] | Timely Response and Containment of 2016 Cholera Outbreak in Northern Zambia | Descriptive analysis of cholera cases in Nsama district between March and April 2016 | 66 cases identified, 8 culture-confirmed, 3 mortalities, 4.5% case fatality rate | Nsama district in Northern province, which has informal settlements along fishing villages, and cross-border transmission from |

|  |  |  |  | DRC |
| --- | --- | --- | --- | --- |
| Gama et al. 2017 [9] | Cholera Outbreak in Chiengi and Nchelenge Fishing Camps, Zambia | Case-control study of risk factors for cholera acquisition amongst fisherfolk in Luapula province compared to controls | 1115 cases in Nchelenge and 257 in Chiengi during the period under review. 76 patients and contracts were subjected to semi-structured interviews assessing their access to WASH interventions such as hand washing soap and running water | Nchelenge and Chiengi districts in Luapula Province |
| Sinyange et al. 2018 [20] | Cholera epidemic Lusaka, Zambia – October 2017 – May 2018 | Description of the outbreak and the interventions done at different stages of the response. And a cross-sectional study on Knowledge, attitudes and practises of households in the affected communities | 5,905 cases and 98 deaths case fatality rate of 1.6 %<br>KAP survey in 98 households | Lusaka, the capital with most cases occurring in Kanyama, a peri-urban unplanned settlement |
| Mwaba et al. | Identification of cholera hotspots | Descriptive analysis using poisson-based | 34,950 participants countrywide, however, | Country-wide, 80% of the cases were Lusaka-based |
| 2020[24] | in Zambia: a spatiotemporal analysis of cholera data from 2008 to 2017 | space and time scan statistics to cover for spatial differences | 29080 of these were in Lusaka over the 10 years |  |
| Mbewe et al 2024 [4] | Survival Analysis of Cholera Patients admitted in Lusaka, Zambia. | Cohort analysis using Cox proportional hazard models to assess survival probabilities and identify predictors of mortality | Medical records for 1529 patients were analysed. overall survival rate was 97%, with the highest mortality being within the first 24 hours of admission | Lusaka |

### Risk Factors and Determinants of Transmission

Male sex, close contact with a cholera case and the use of borehole water were found to be risk factors for cholera infection [19–22]. Drinking water sources were found to have inadequately low free-residual chlorine (FRC) in up to 71% of households surveyed [20,23]. Thirty-one per cent of those households with inadequate FRC had evidence of faecal contamination. Low latrine coverage, poor drainage systems, and sharing latrines [8,19] were also documented vulnerability factors that allowed for the perennial occurrence of cholera in some localities, particularly unplanned settlements such as the fishing villages in many of the areas bordering lakes [9,24]; and in peri-urban areas with high population densities such as Lusaka and the cities on the Copperbelt [20,24]. Whilst poor hygiene practices (mostly superimposed on people due to lack of facilities) were a notable risk factor, consumption of food products, particularly fresh fish was not associated with an increased risk [13].

The availability of drinking water and the quality of the peri-urban areas of Lusaka was assessed. It was found that in areas underserved by the municipal utility companies, private borehole companies known as ‘Water Trusts’ would operate small shops known as ‘kiosks’ where community members could go and draw small quantities of water in buckets at a minimal cost to cover the fees only, and not for profit [23]. These trusts treated and provided water to the communities in these water-stressed unplanned settlements as an adjunct to the provincial utility company, and yet they were found to serve less than 60% of the communities in need of their services [8,19]. Despite this limitation, they were noted to present a safer alternative than privately consumer-owned boreholes and shallow wells in terms of faecal contamination with *Escherichia coli* and nitrite content of the water [23]. Those unable to afford the kiosk water would tend to use unsafe surface water sources such as shallow wells in their locality [19,20]. These presented the highest risk of contamination particularly due to topographical features such as the high-water table in Lusaka Province leading to a high risk of contamination of these shallow wells from nearby pit latrines [22,23].

The smallest surveillance unit of population reported was the ward level. It was found that the greatest risk for cholera was found in the wards with the densest populations, unimproved sanitation and evidence of *E. coli* contamination of piped sources [11]. Elsewhere, as was seen in Kabwe, through an environmental sampling of groundwater using polymerase chain reactions (PCR) tracers, there was evidence of groundwater contamination with documented environmental vibrioni [22]. The authors postulated that private boreholes are vulnerable to contamination possibly due to incompetent casing which may then provide an artificial pathway for the vibriones from contaminated ground sources and hence pose an even greater risk. Supporting this, was the rapid decrease in cases seen during outbreaks, when there was an increased provision in water and sanitation services such as hyper-chlorination of the water utility lines, provision of safe water through emergency tanks in the hotspot areas [25] and with the use of reactive OCV campaigns [26].

### Inter-district and inter-country spread of outbreaks

Risk factors for continued outbreaks between the peak years included increased poverty and inadequacies of social services due to rural-urban migration [8,19,27]. Similarly, movement between neighbouring districts (24) and neighbouring countries [16,28,29] were identified as factors associated with epidemic cholera in Zambia. Chirabombo and colleagues documented how naïve districts neighbouring traditional hotspots such as Lusaka, can also then present with outbreaks of their own with evidence of local transmission [12]. The question of environmental persistence versus reintroduction into the district from neighbouring countries such as the DRC and Tanzania, which equally have continuous outbreaks, has been documented [9,24,30]. This is reaffirmed by laboratory studies and descriptive analyses of genomic sequencing isolates that showed a wide genetic diversity [16] and close linkage with isolates from other parts of the Great Lakes region [28,31]. This underscores the need for both a decentralised approach at the district and ward levels but also shows a need for enhanced cross-border surveillance and possible cross-border joint responses [31,32].

### Clinical Characteristics and Host Predisposition

Globally, there is a dearth of information on the clinical characteristics of patients affected by outbreaks beyond general case counts and case fatality rates [33]. Little is known about the proportion of pregnant women, elderly or paediatric patients affected by cholera, nor the number of patients presenting with co-morbid conditions or other complications of care. What was seen is that having received limited education and being older than 55 years constituted one risk factor for increased mortality [34]. There was a slightly higher proportion of patients documented to have died prior to arrival at the treatment facilities (i.e. at home or community deaths) vs in the facility (55% community deaths vs 45% facility deaths, respectively); with similar proportions having been noted to have received some oral rehydration solutions (ORS) before presentation or death. Intravenous fluids were not available beyond Cholera Treatment Centres (CTC), and there was no documented use of Community Oral Rehydration Points before the 2023/2024 outbreak. Prior antibiotic use was not found to be protective, although noted that patients often took metronidazole which is not one of the recommended agents [34]. Adequate ORS was protective [34], yet it was clear that there were disparities in the availability of ORS, particularly in rural communities [35]. In younger patients, cholera was noted to be an important cause of morbidity and mortality in the under-five age group, with increasing antimicrobial resistance over the years [32,36]. Case management was reported to have improved, with reductions in the case fatality rate (CFR) decreasing from 6.7% in 2000 to 1.7% in 2010 [8]. However, as of the 2018 outbreak, the case fatality rate hovered around 2.5% [20]. The case fatality rate of the 2023/2024 outbreak was 1.3%, with increased documentation of community deaths [3]. For inpatient fatalities, there were higher odds of dying for those with pre-existing comorbid conditions [4].

### Vaccine availability and effectiveness

Vaccines are known to be a useful tool for community-level interventions for controlling waterborne diseases such as cholera, in places where access to water, sanitation and hygiene remains limited [37]. More recent studies have used the new Euvichol plus®, which is the Eubiologics® bivalent vaccine of El Tor and Ogawa presented in glass containers as opposed to plastic vials to improve cold chain in humanitarian crises [42]. They have shown a higher vaccine efficacy in the two-dose strategy than the single dose (at 74% and 81%, respectively [43]; and that reported OCV administrative coverage is often much lower than the actual coverage which was found to be 66% of people getting both doses, which may further lower efficacy rates [44]. Questions persist about the very high dropout rate of 18% between the two doses [44]. Similarly, it remains to be seen the effect of previous preventative campaigns, as a lead up to future multiyear preventative vaccination campaigns.

Pugliese-Garcia and colleagues attempted to explore the factors influencing vaccine acceptance and hesitancy in the hotspot districts of Lusaka. They found that traditional remedies, religious beliefs and alcohol use persist as impediments [45], as does a background mistrust towards Western medicine [46]. There was an overarching sense of helplessness or ‘fate’ as the participants were aware they could not change their living conditions and did not realise their ability to use safer water practices to protect themselves [46].

Investigation of the immunogenicity of the vaccines in a controlled population in one of the high-risk fishing villages found no significant difference in vibriocidal antibodies at two weeks or six months and provided evidence for the delayed dosing schedule [47], but also waning immunity beyond 12 months [48]. The group found no influence of ABO blood groupings on vaccine response [49]. HIV-positivity was found to reduce immunogenicity in these individuals regardless of the CD4 count, whilst serum vitamin A levels had no effect positive or negative [50]. Elsewhere, there was a suggestion of vitamin A supplementation as a possible adjuvant to improve T-cell expression following vaccination, particularly in children [51,52], which may offer a gateway into host-specific factors for improved immunity and transmission dynamics. There was no work yet published on the role of the gut microbiome in cholera vaccine responsiveness nor protection in the face of household exposure. Most recently a comparison of vibriocidal antibodies in naturally infected vs vaccinated individuals was found to be comparable, with peak immunity seen around day 19 post-infection and waning after day 30-39 [53]. The authors suggested the need for booster vaccinations, particularly in high-risk areas as a possible public health protective strategy.

The use of a single-dose campaign of Sanchol® was found to be cost-effective, amounting to just under $ 1 million to vaccinate 500,000 people [26]. A further evaluation of the cost of cholera illness and the cost-effectiveness of the single-dose campaign in Lusaka was close to $1000 per disability-adjusted-life year (DALY) averted, especially in those above the age of 15 years [54]. The social implications for affected communities have not been deeply studied, nor the cost-benefit analysis of community-based interventions and health education initiatives in the hotspot districts. With the increasing size of the outbreaks, it remains to be seen the cost-effectiveness of reactive campaigns, and also the macro-economic effects of the overall cholera responses.

### Climate variability

The role of climate variability and extreme weather events cannot be ignored with a strong association between the onset of rainfall and epidemic outbreaks [20]. Cholera outbreaks in Zambia, like many other African countries, are seasonal [22,30], differing from the Ganges Delta where it occurs perennially [30]. The outbreaks start with the onset of the rainy season in 71% of cases and have been associated with 50% of all recorded drought years. Outbreaks are expected to increase in frequency by 300% in the near future with recurrent El Nino events [55]. Following seasonal rains, the larger outbreaks are often heralded by flooding which is a specific sequel of torrential rains possibly enhanced by climate change. Flooding has been associated with damage to WASH infrastructure, and the decay of flooding countermeasures such as clogged-up drainage canals and sealing of ground passages for water, particularly when big cities such as Lusaka are afflicted [20,55] further compounding the problem. Reduced rainfall (i.e. drought periods) may also increase cholera outbreaks as seen in the U-shaped occurrence of diarrhoeagenic bacteria such as *V. cholerae* with rainfall and pathogen proliferation [56]. Groundwater drilling during the drought years, if not carefully planned, will worsen the already water-stressed situation in certain parts of the country [55]. The anticipated periods of droughts in the near future are expected to exacerbate rural-urban migration into the peri-urban slums, further compounding the water-stressed situations and the likelihood of larger cholera outbreaks [55].

Mathematical modelling was used to predict the expected time to extinction of cholera in Lusaka and based on previous estimates of a second wave in each outbreak found that heavy rains were associated with an increased environment to human transmission [27]. They warned that environmental vibriones could persevere for eight months to six years in the environment especially the shallow wells and areas with poor drainage, hence future outbreaks would be longer and more severe. They also recommended enforcement of the multisectoral cholera elimination plan, which sought the combination of WASH interventions with periodic oral cholera vaccinations [27]. All the reviewed articles are listed in Table 2 with their key findings and possible mitigating factors that can contribute to cholera control and elimination in Zambia.

**Table 2:**
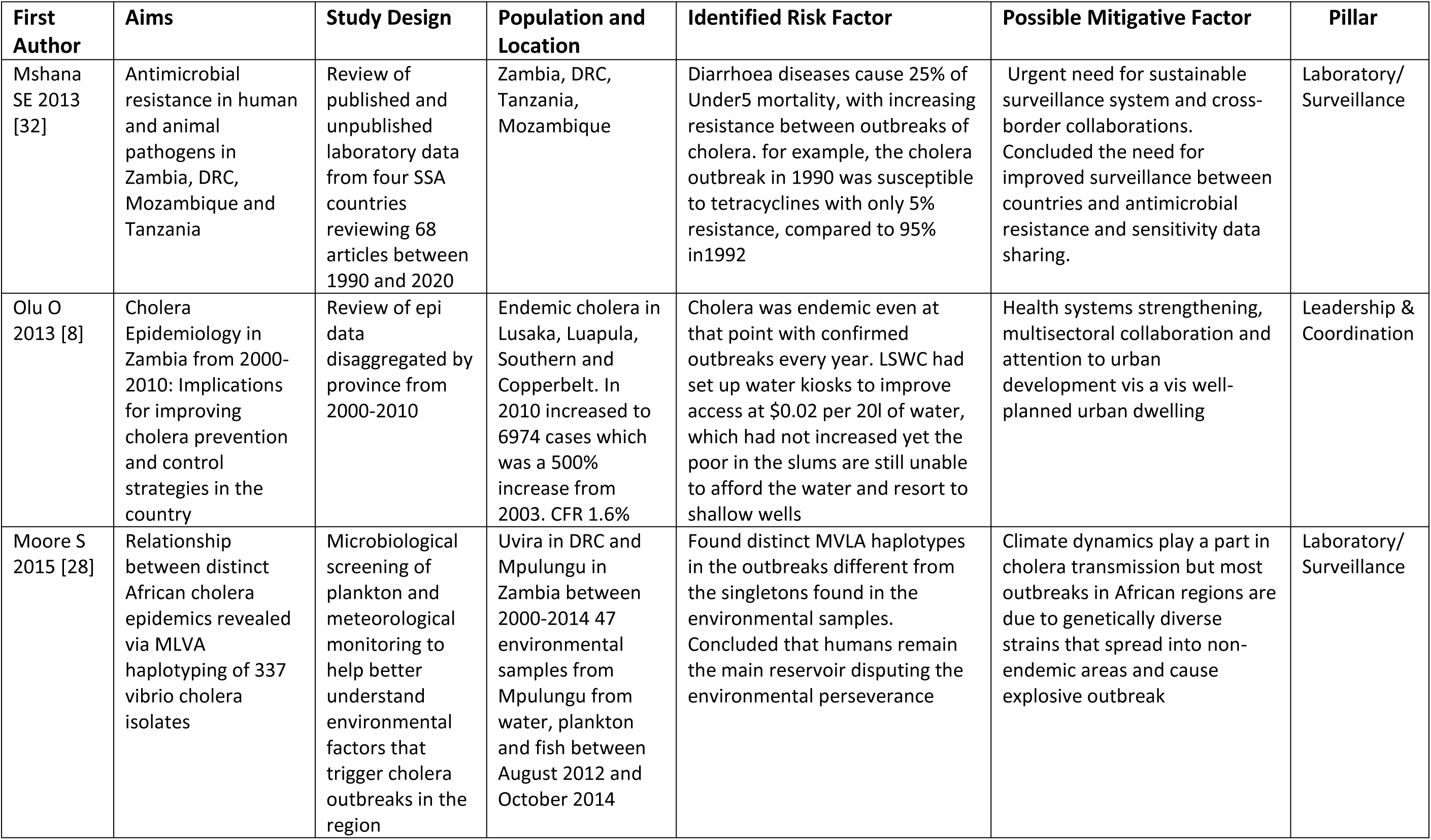
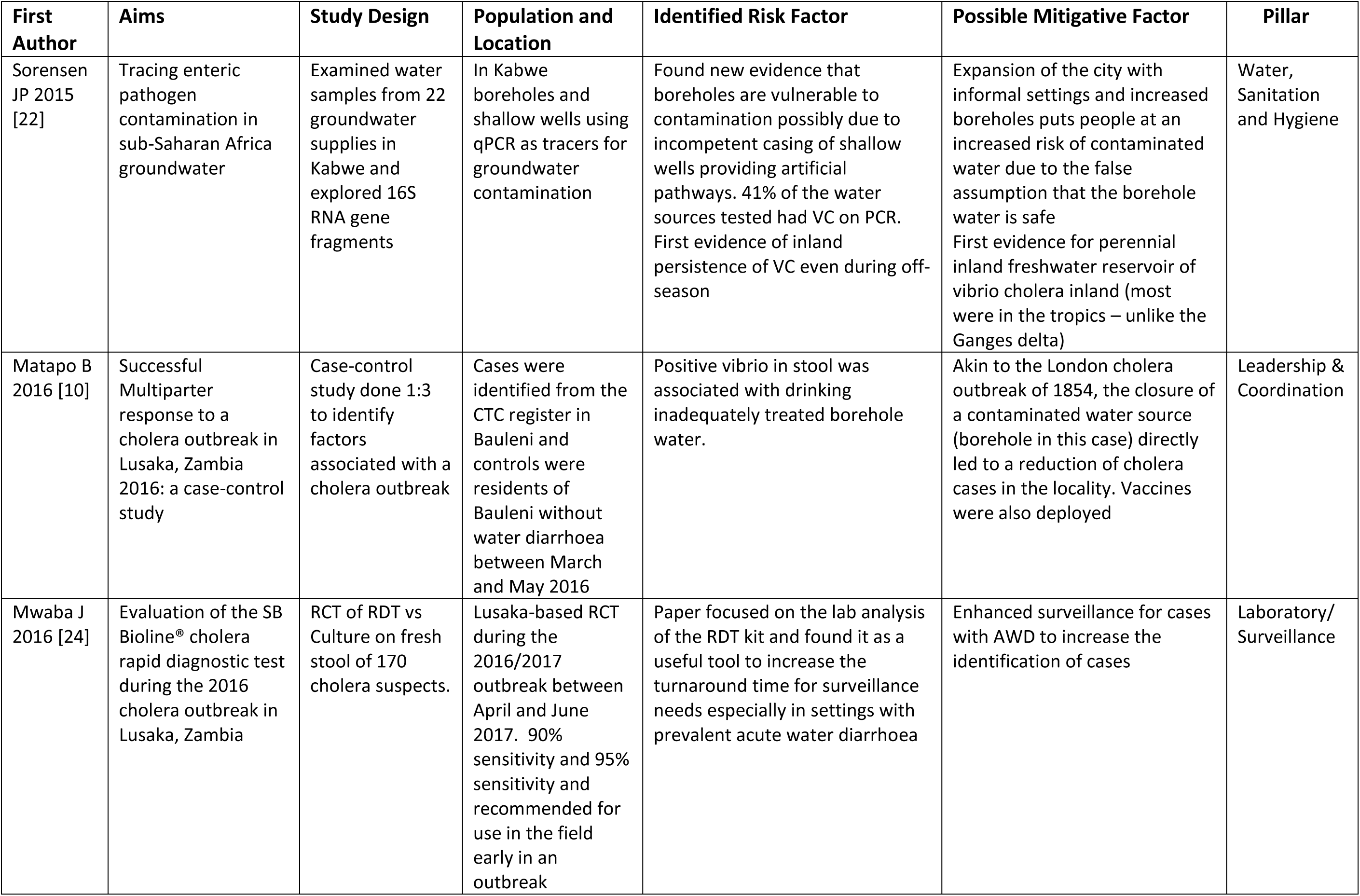
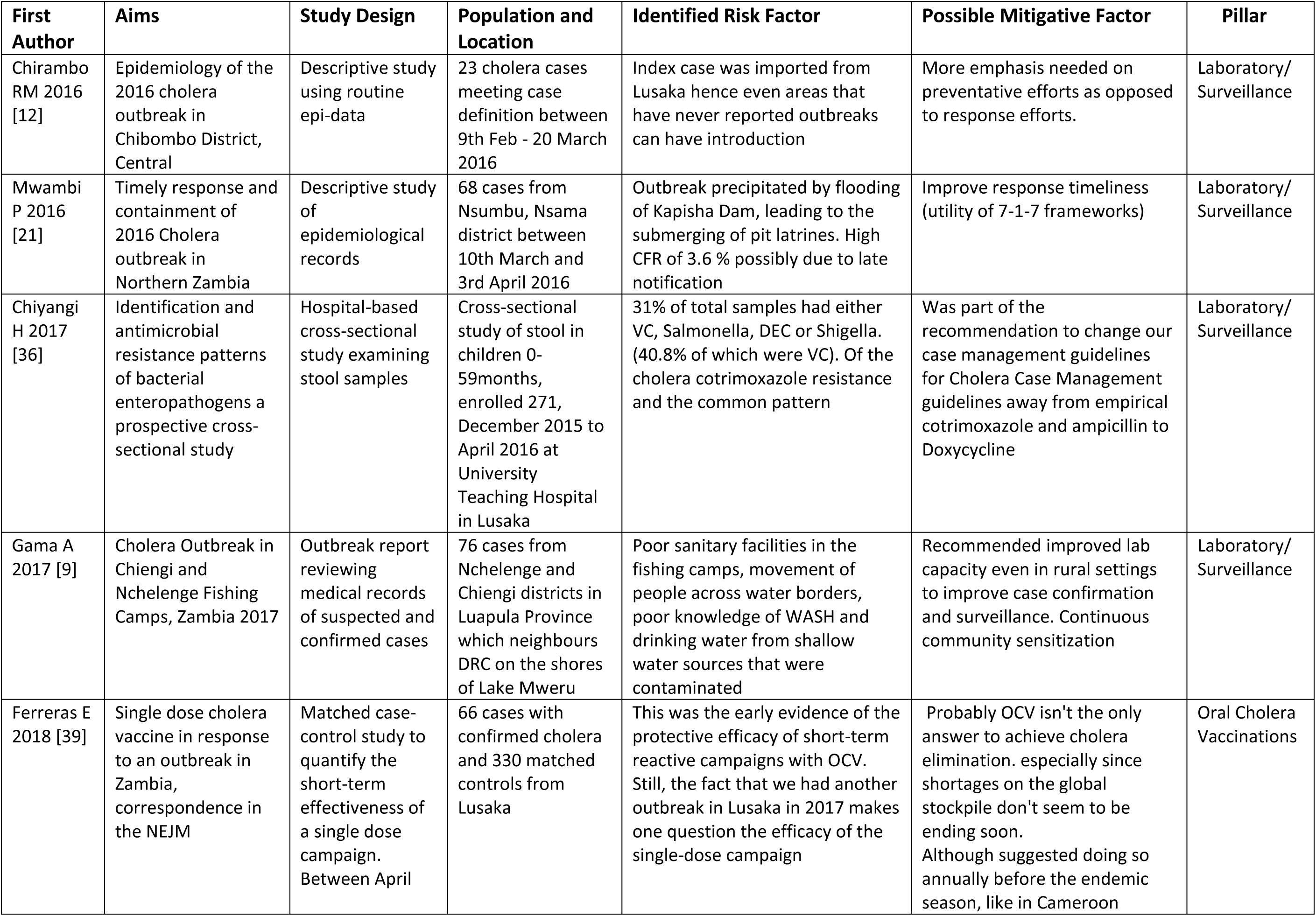
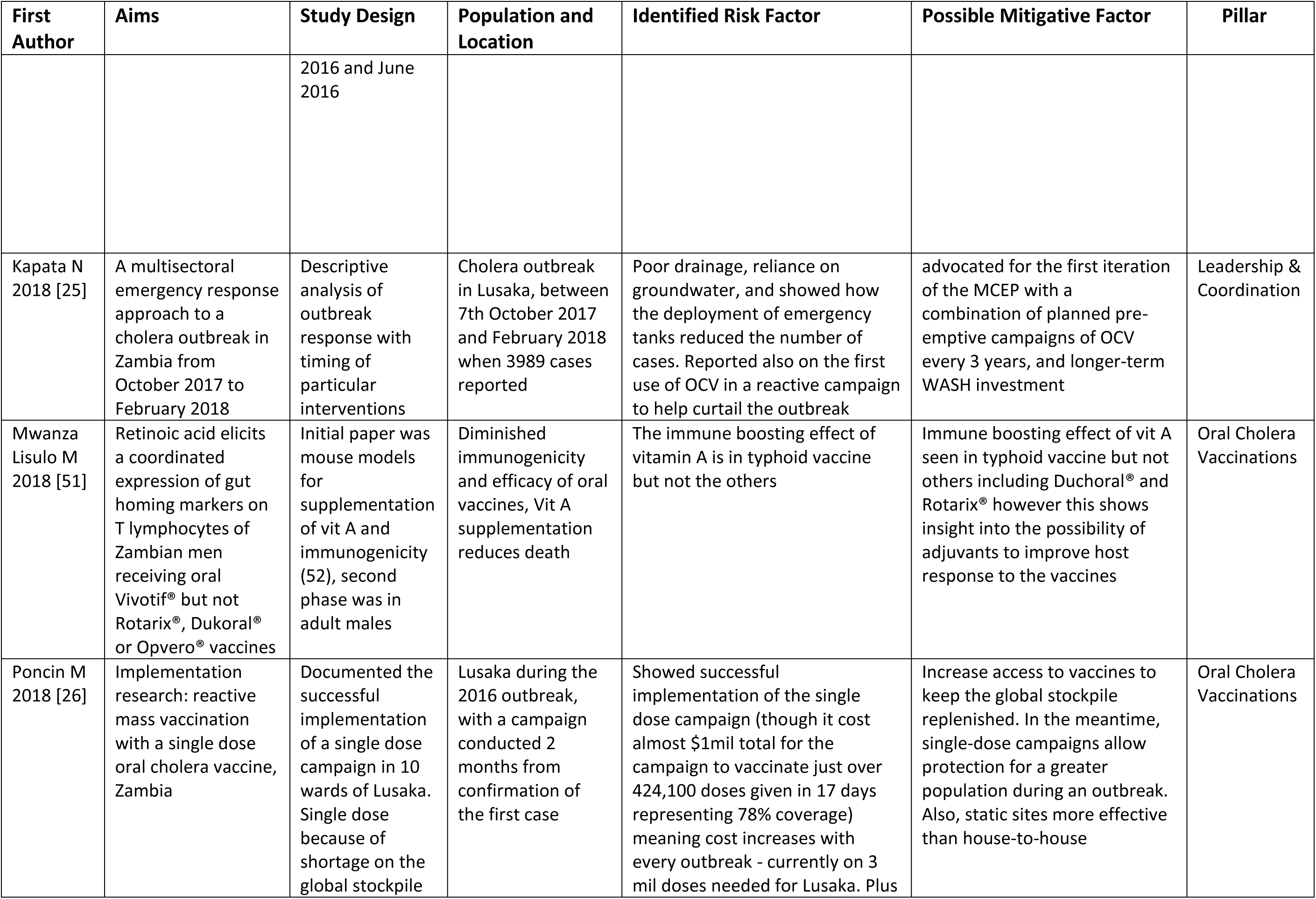
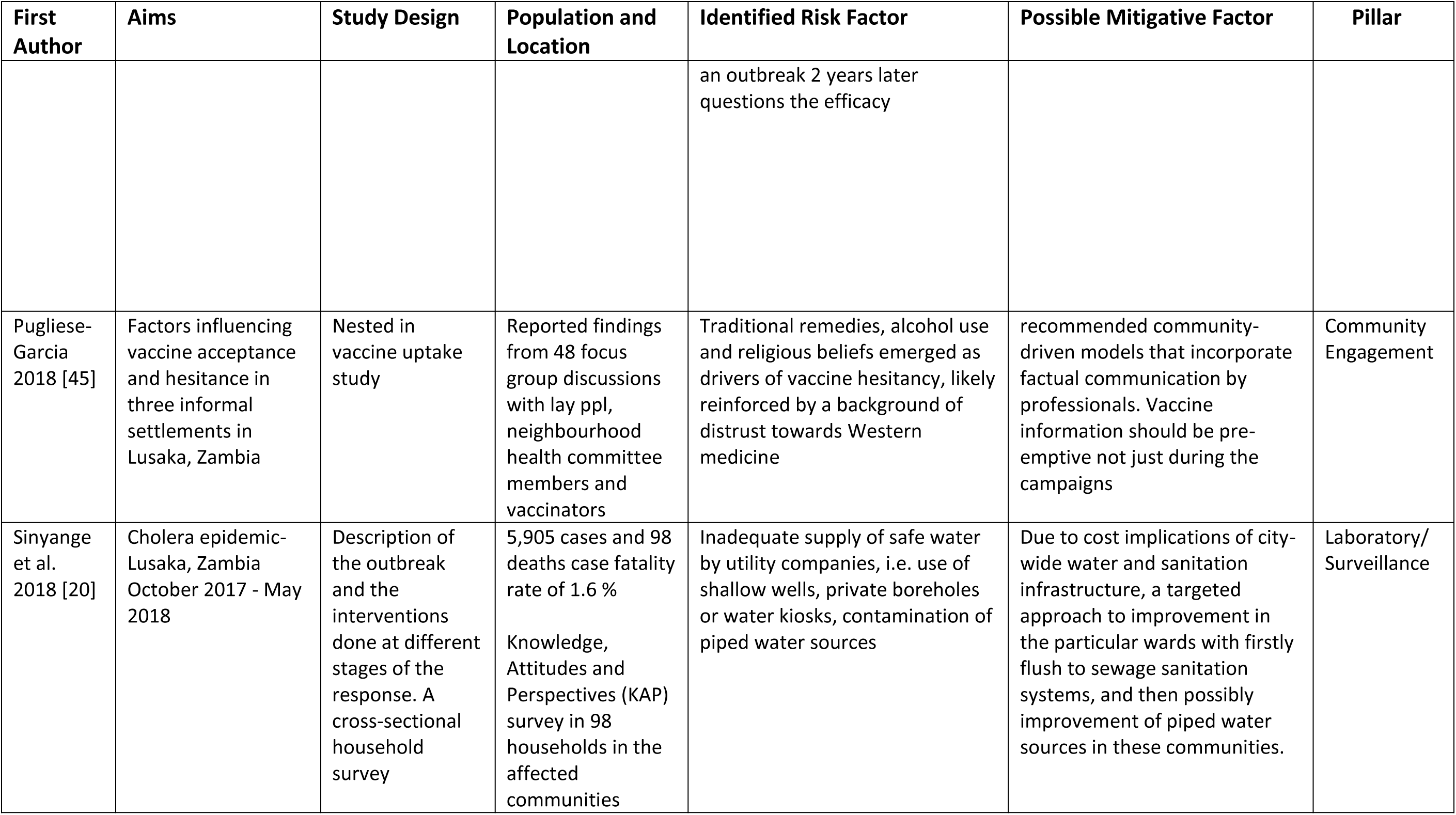
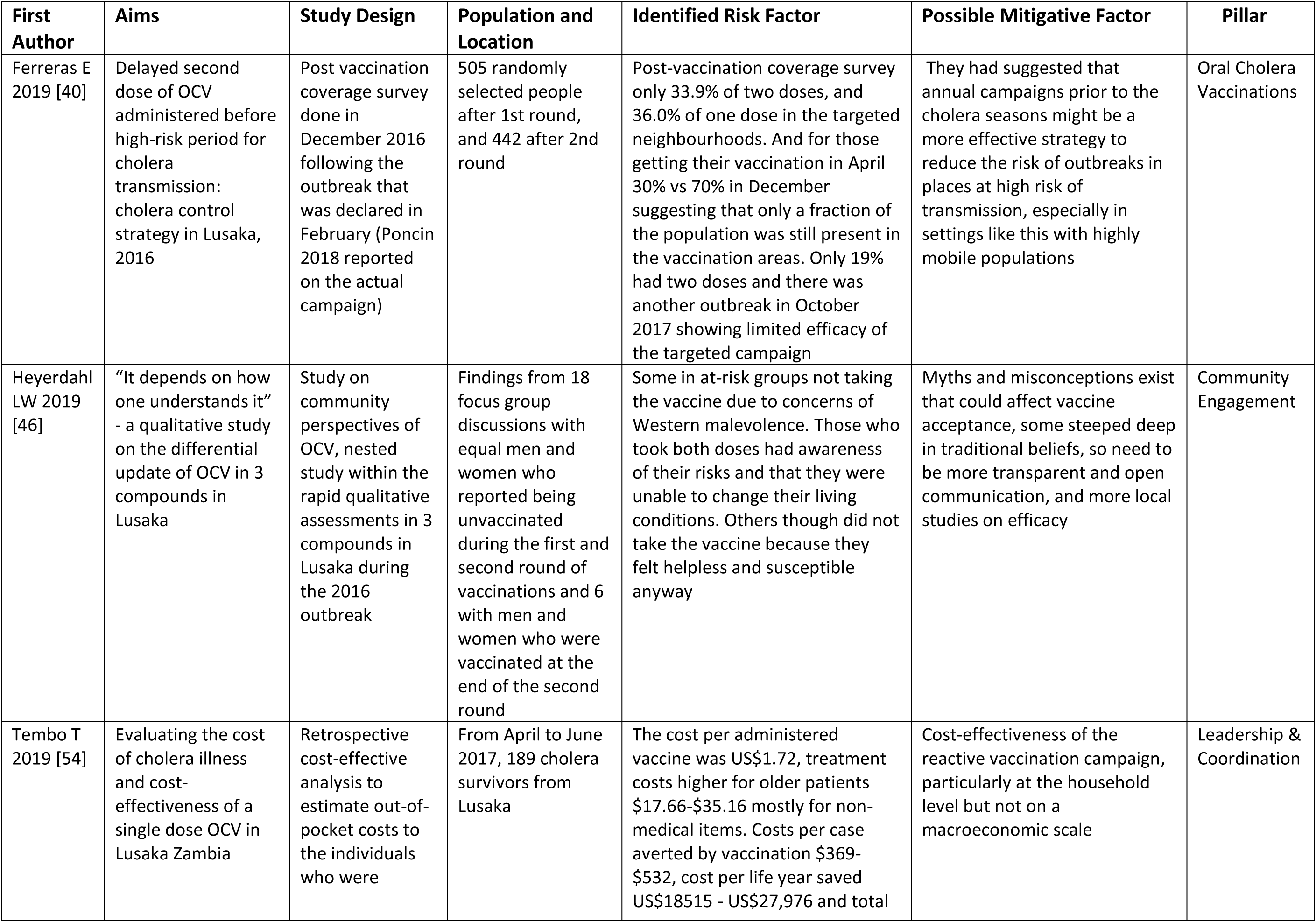
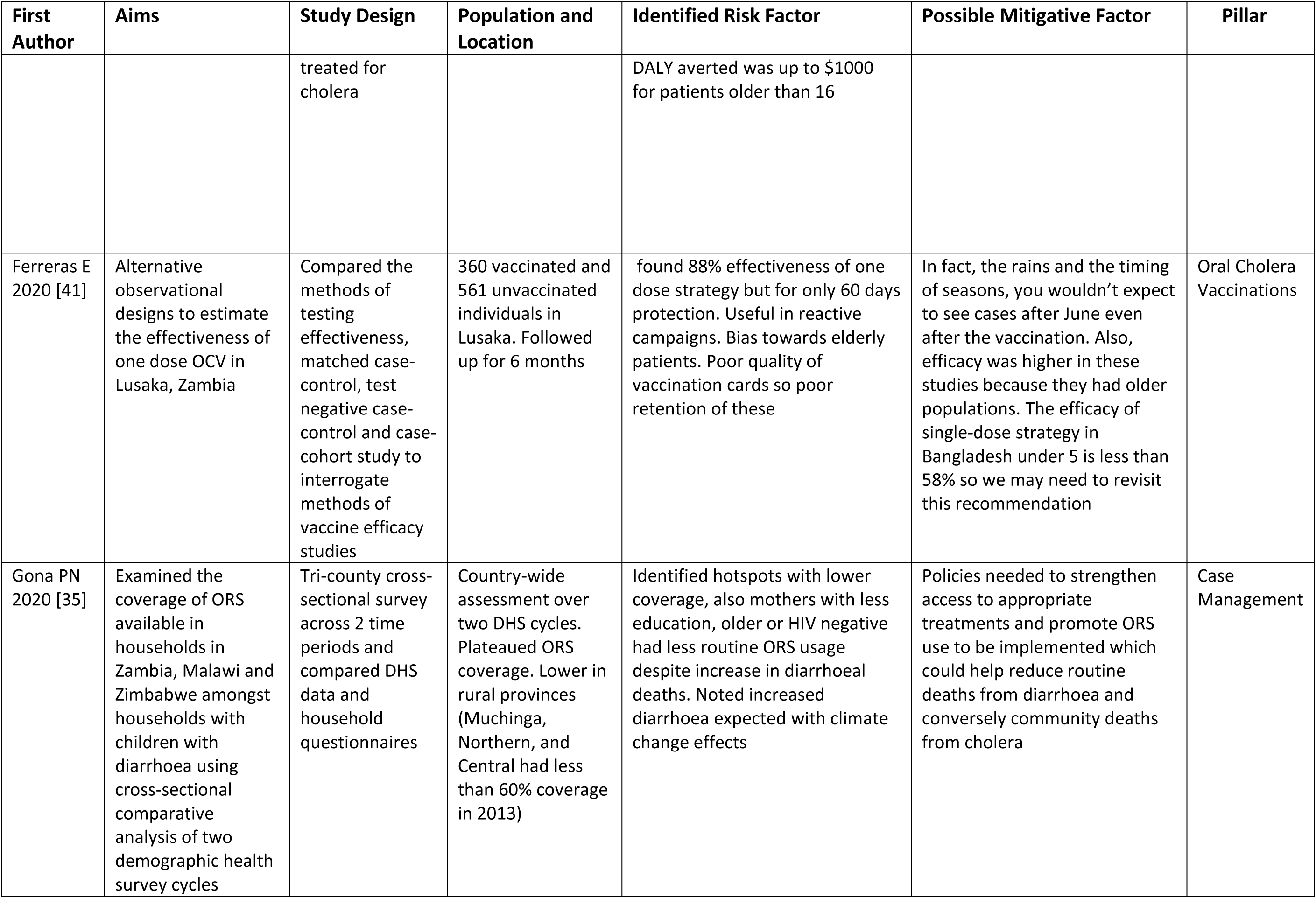
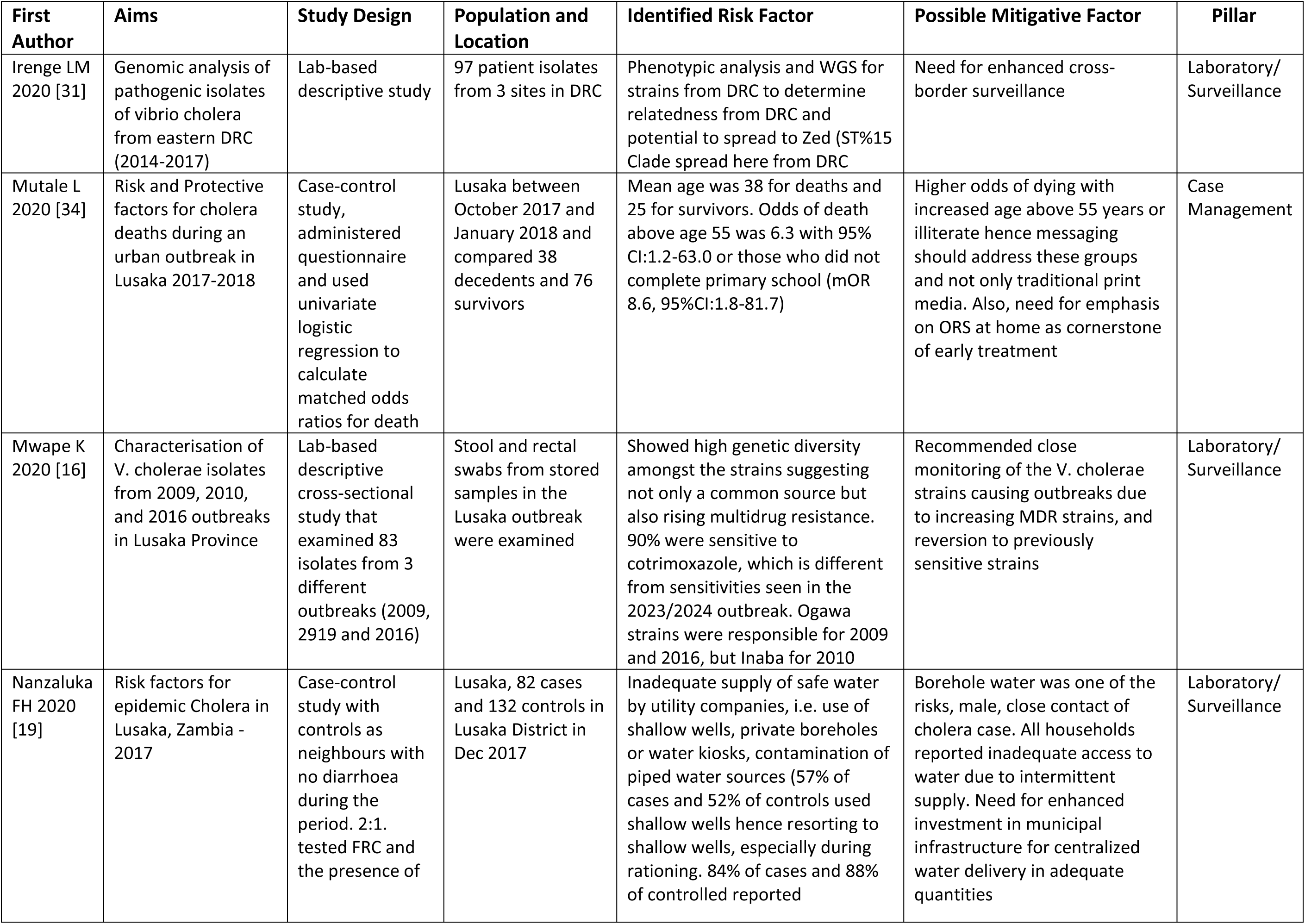
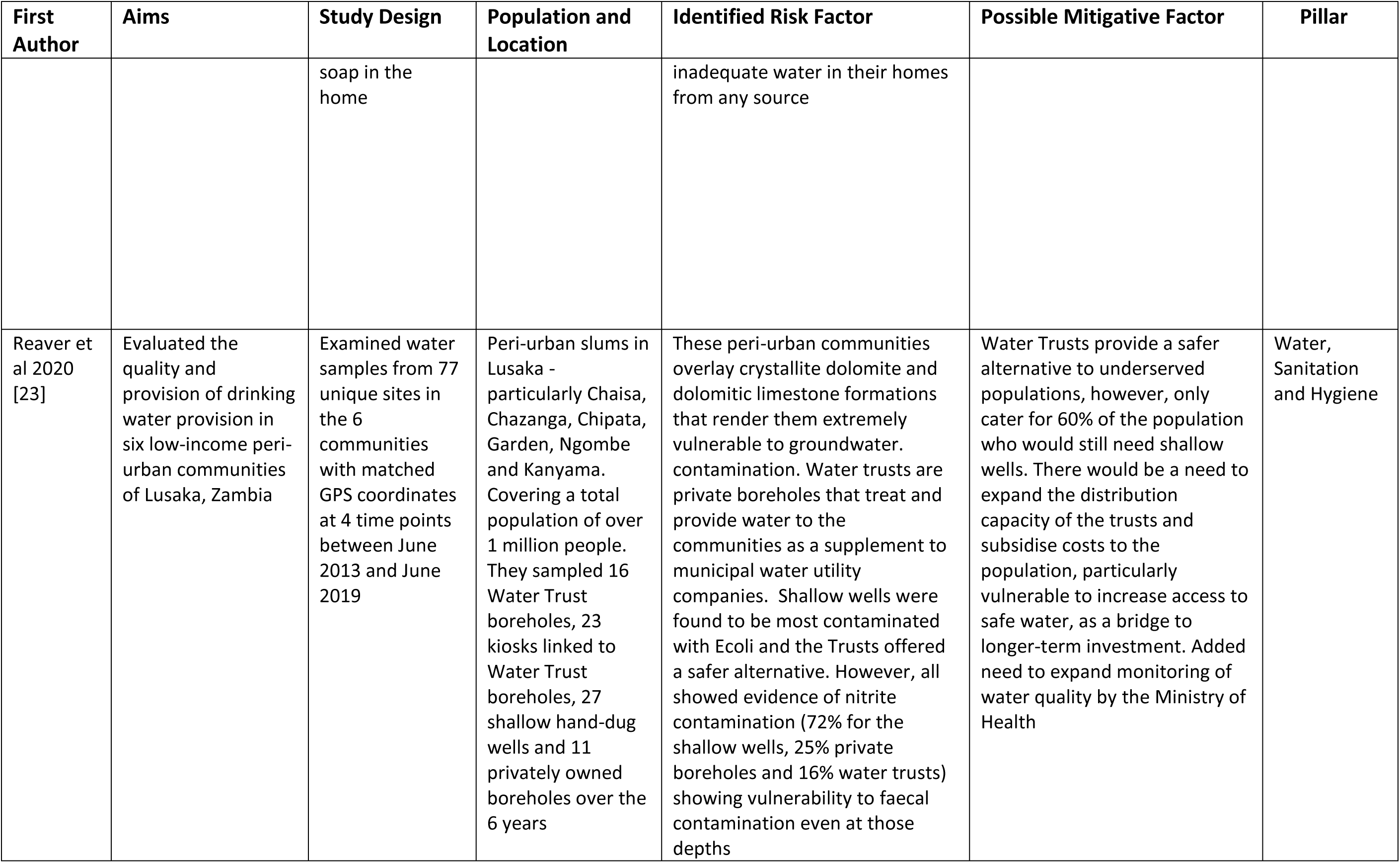
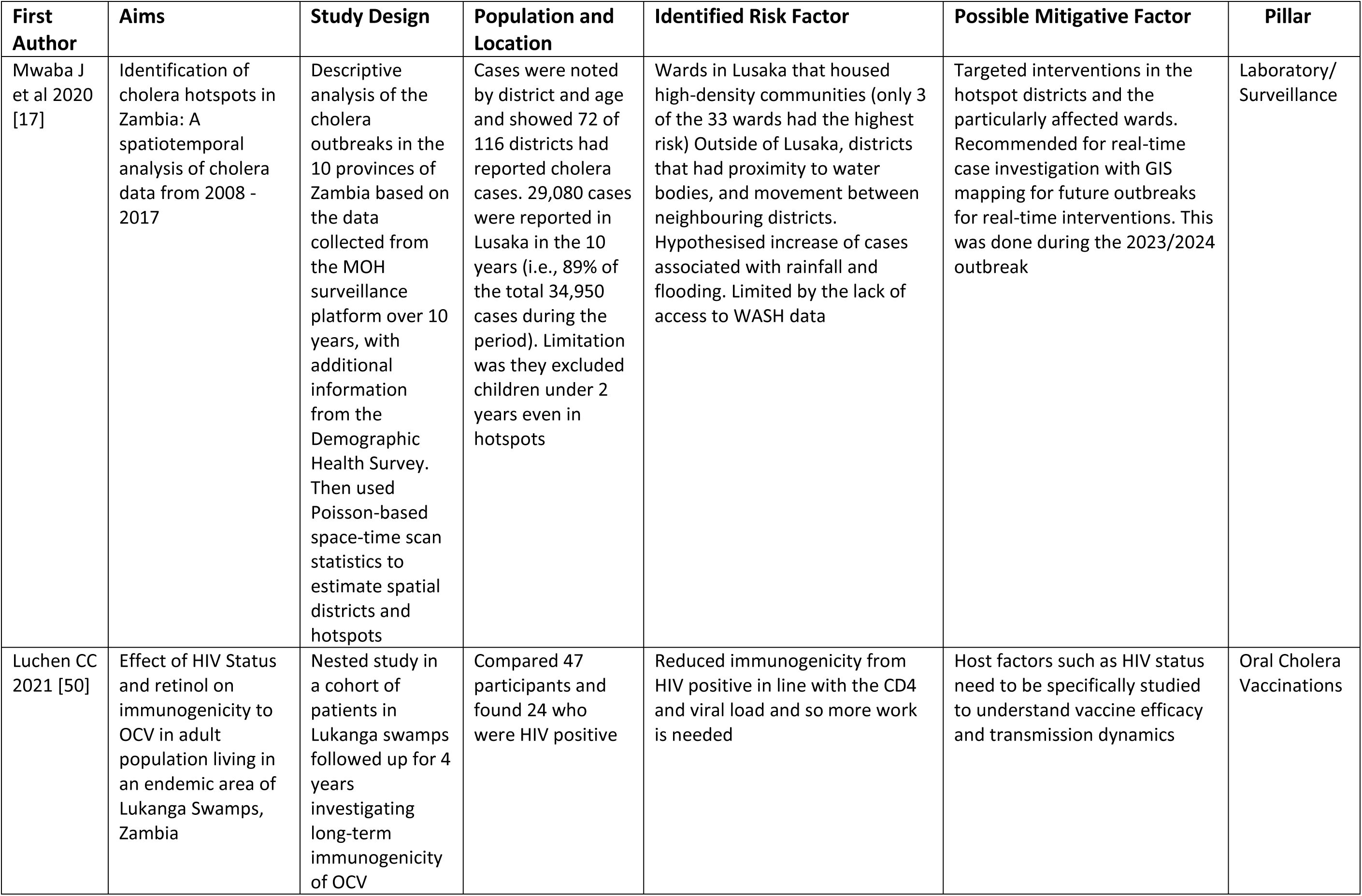
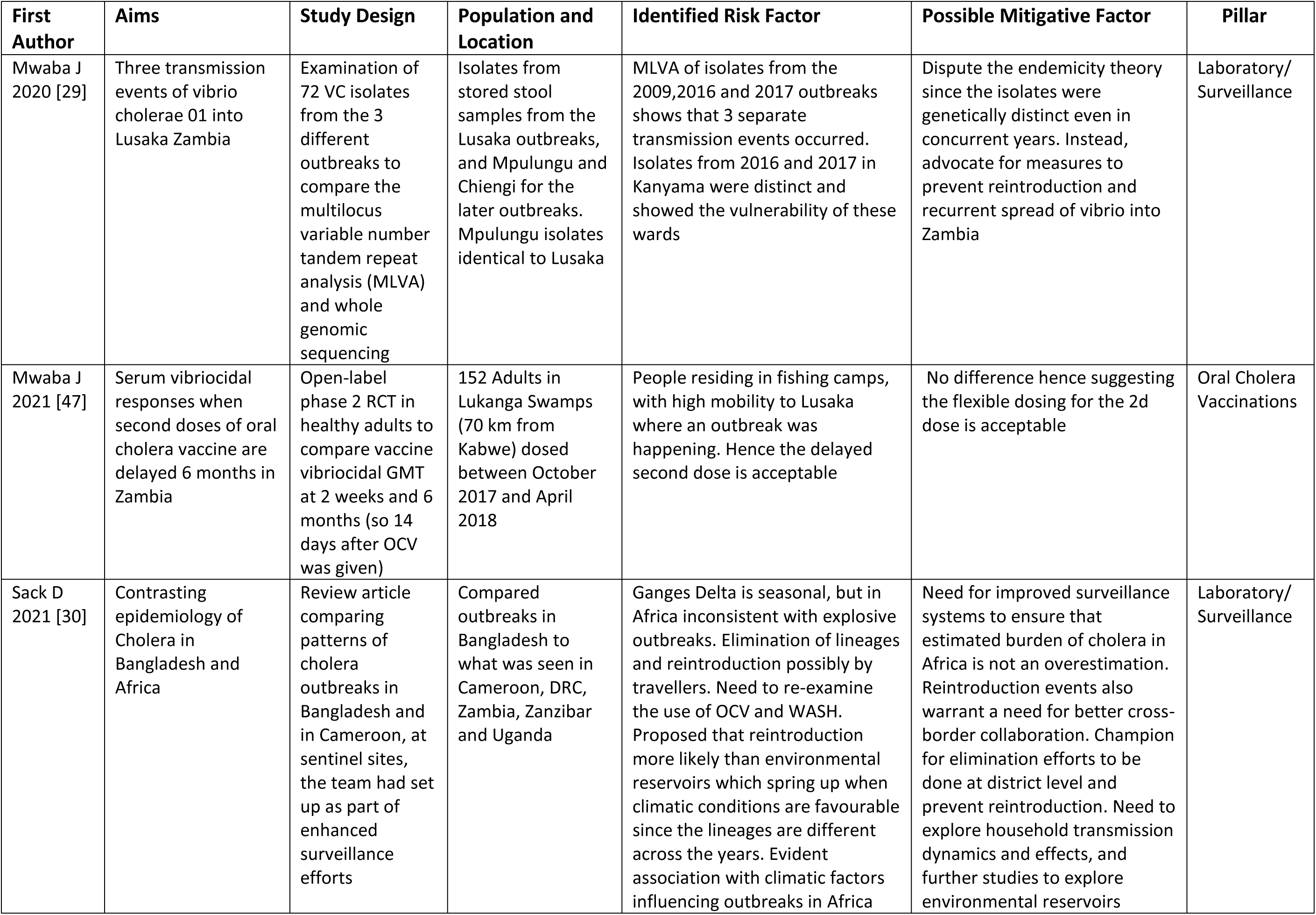
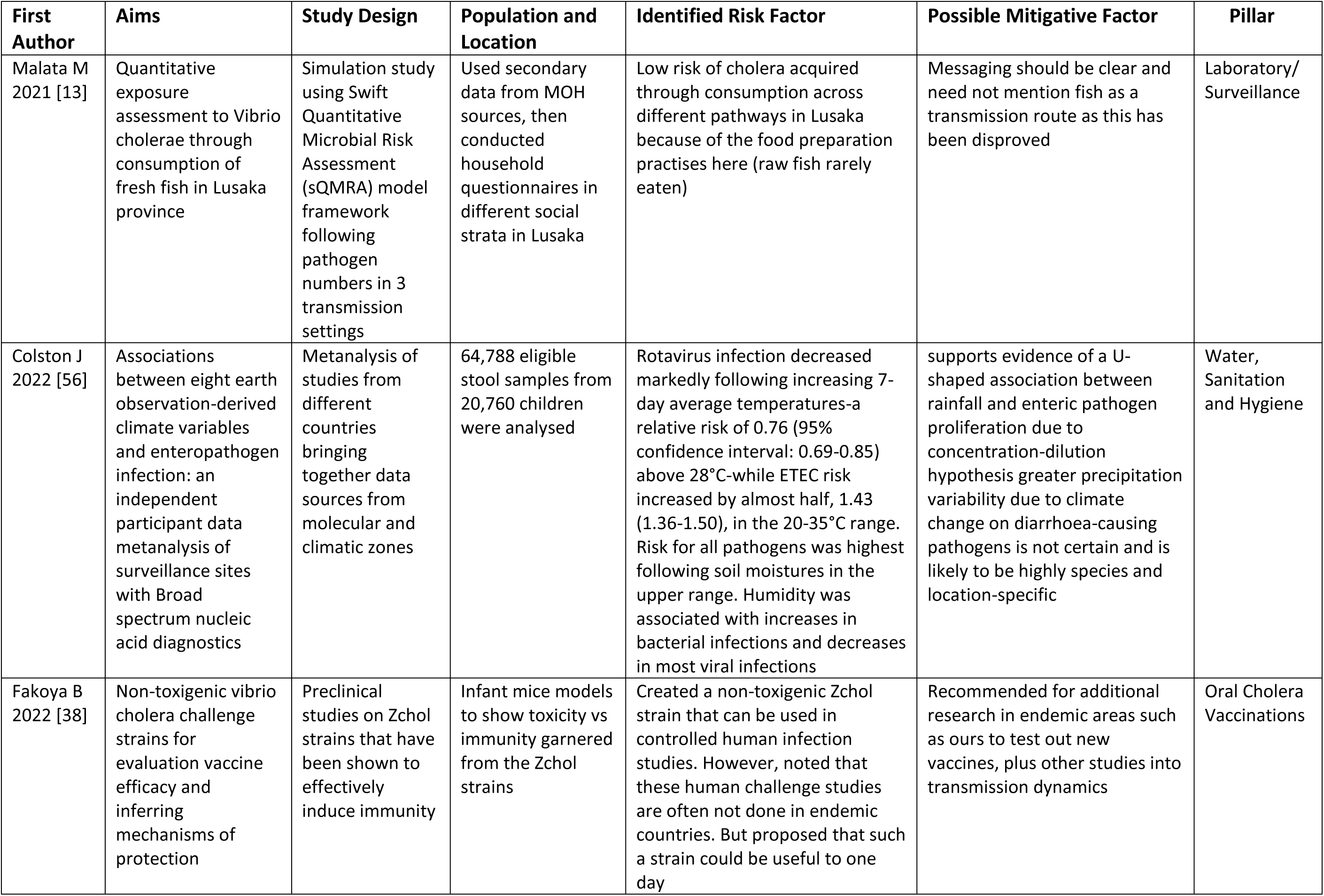
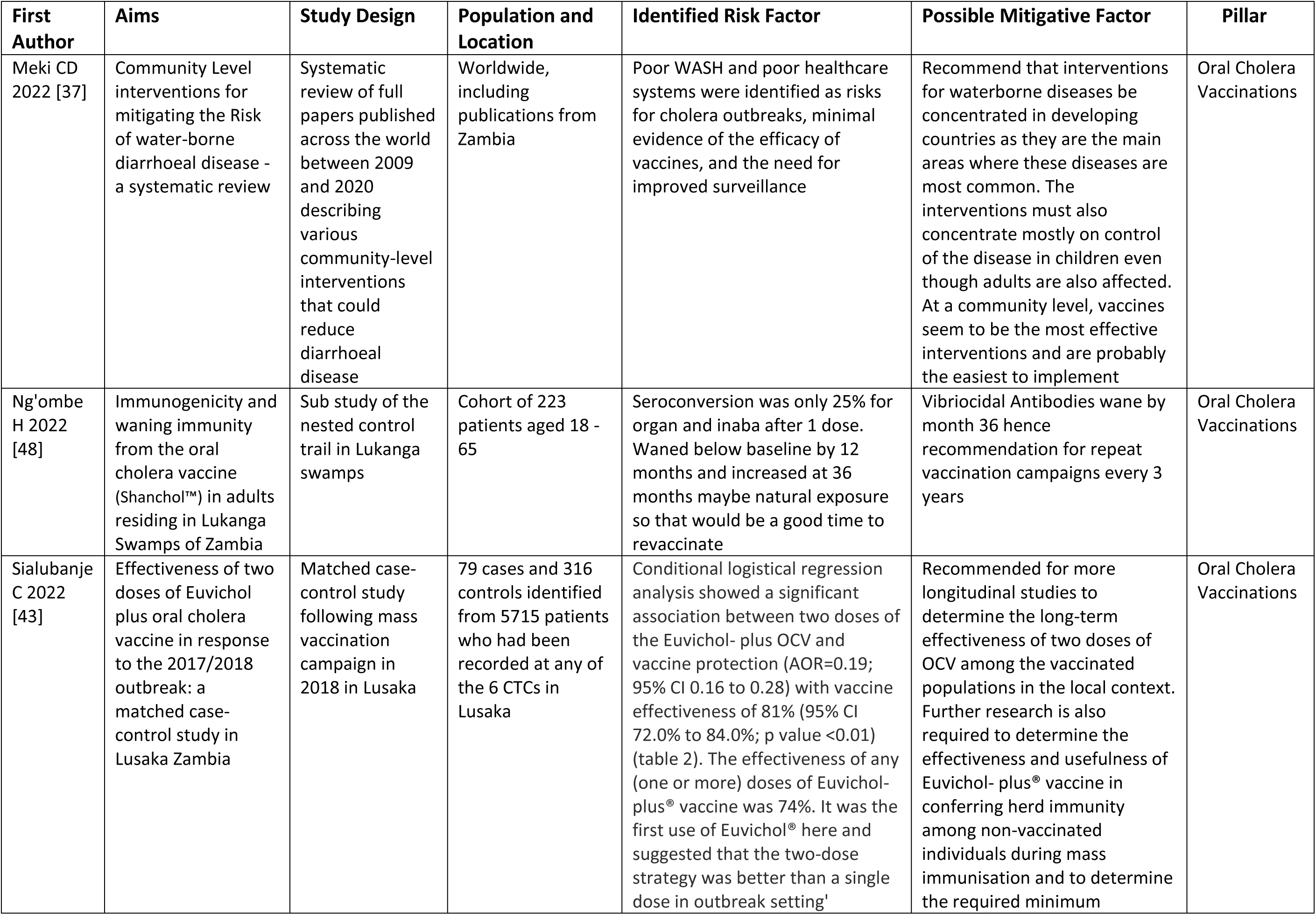
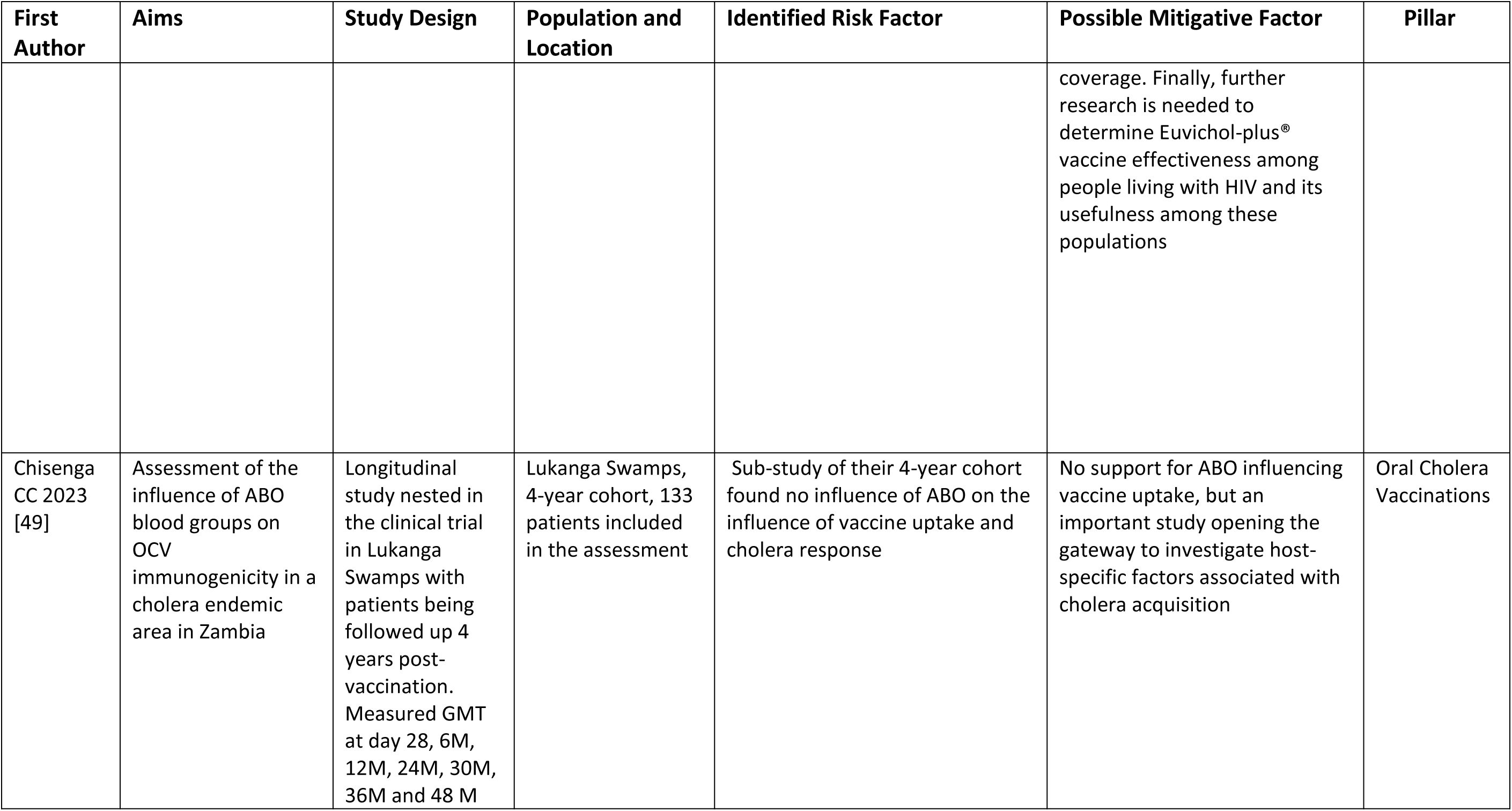
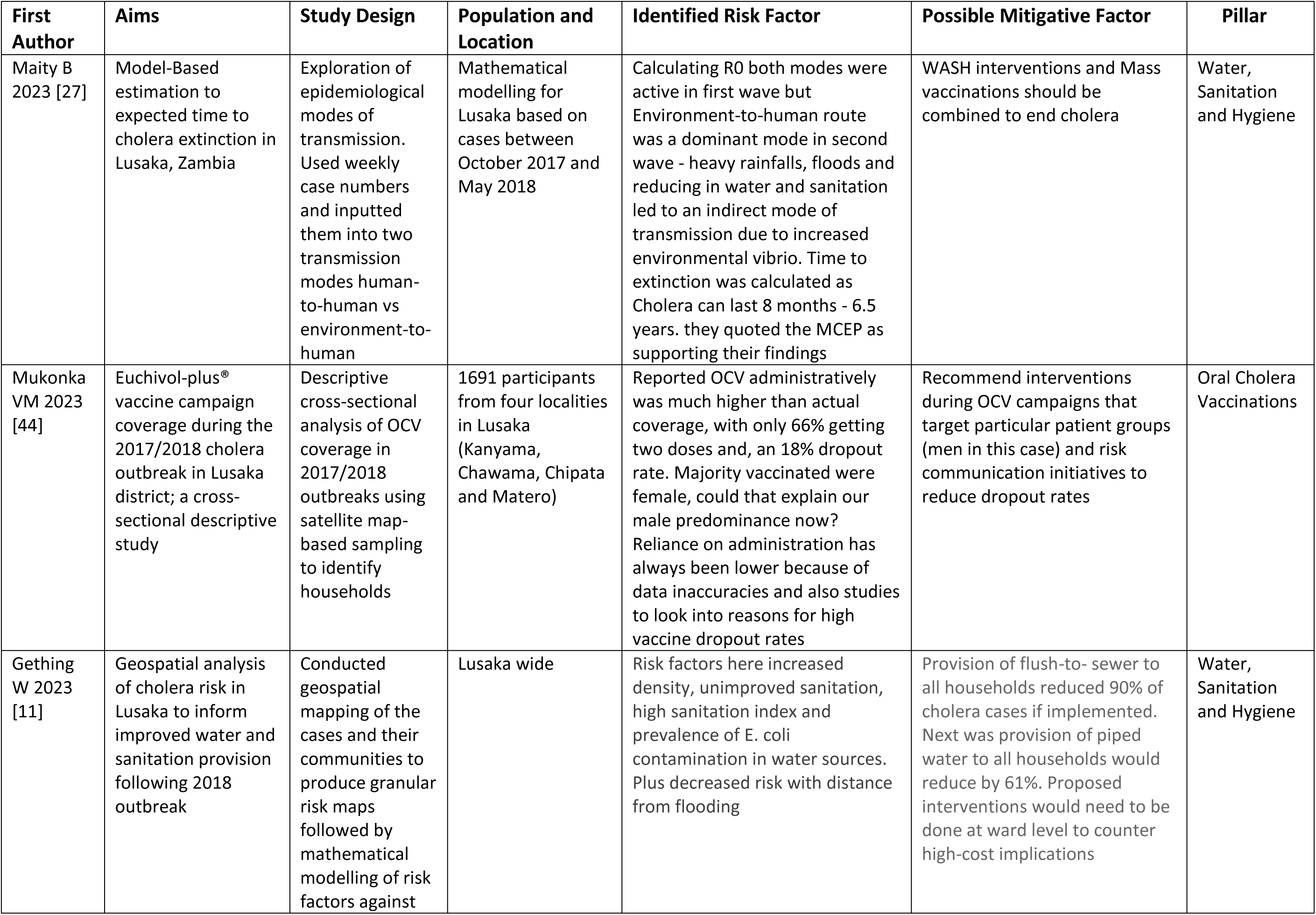
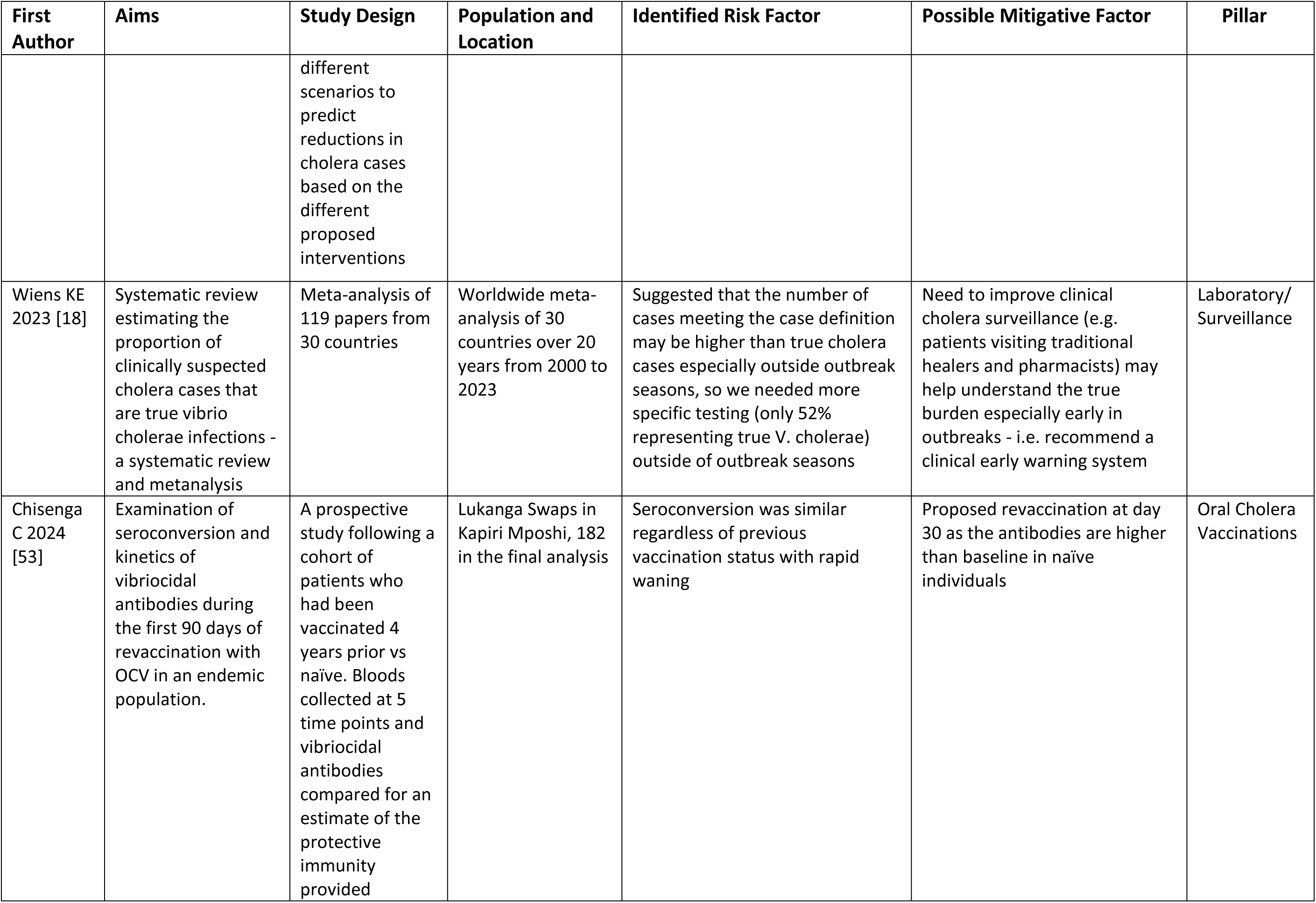
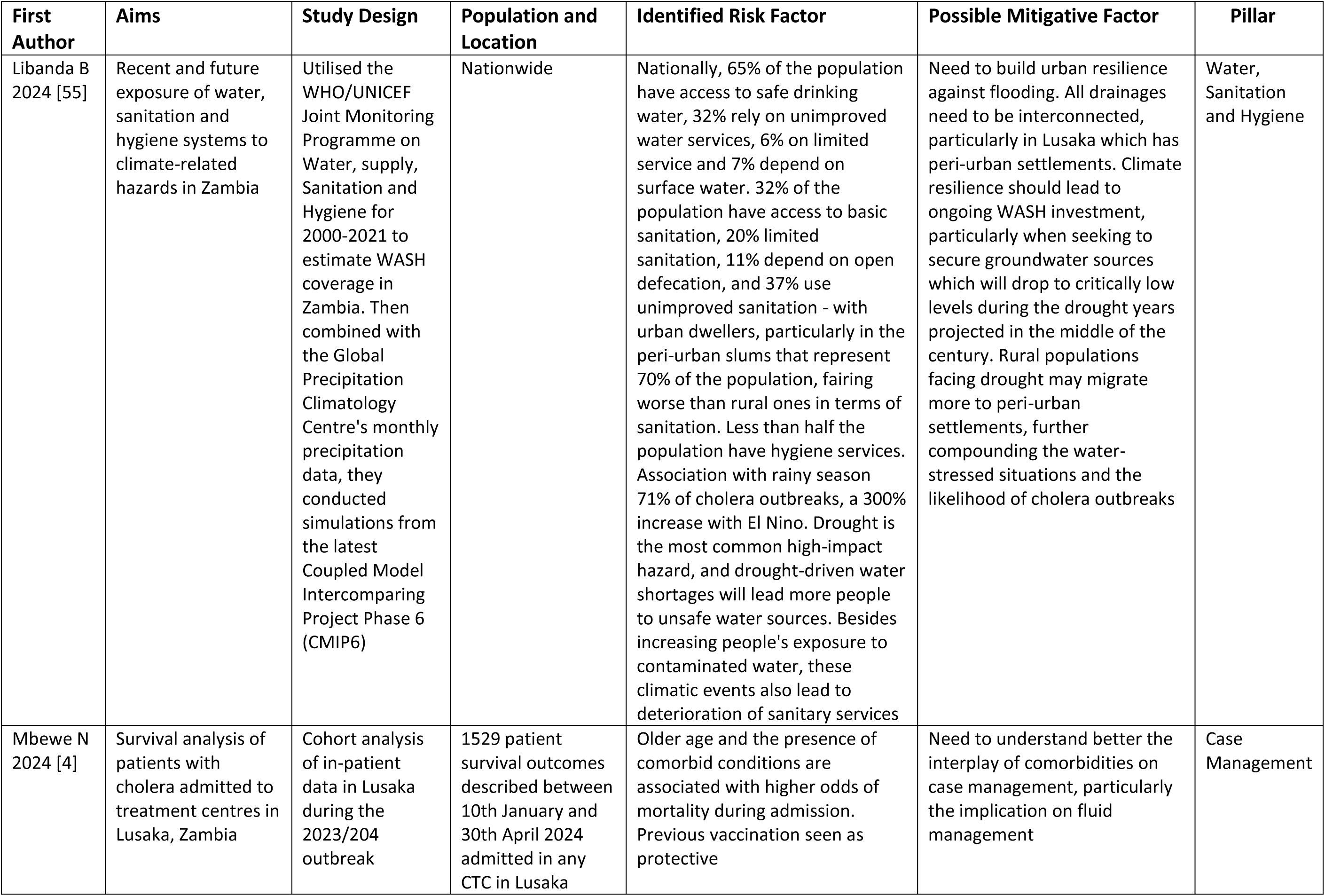
Comprehensive Review of Cholera Research in Zambia (2013-2024): Aims, Study Design, Population, Identified Risk Factors and Mitigative Measures.

## Discussion

Since the first documented outbreak in 1977, Zambia has recorded major outbreaks every three to five years with increasing intensity and fatality [4,8,11]. The outbreaks were predictable concerning the timing in the calendar year and with an increasing frequency related to climatic conditions and urbanization [11,20,23]. Because most of the reporting is done based on case definitions during outbreaks, it is postulated that the true burden of cholera in Zambia, like other parts of the world, is underreported outside of explosive outbreaks [57,58]. The major risk factors for recurrent outbreaks in the country were poor access to water and sanitation services in urban unplanned settlements and the rural fishing villages [9,19,24]. These factors were found persistent even in the 2023/2024 outbreak, which is the largest to date [4], different from other cholera-prone areas which are often coastal areas in South Asia [30] or places with humanitarian crises and conflicts such as Northern Nigeria and Haiti [59,60].

Zambia was not considered endemic of cholera at the time of the development of the first multisectoral cholera elimination plan (MCEP). However, the increased frequency and almost annual occurrence of outbreaks in certain localities necessitate its reclassification as an endemic country eligible for cholera control as opposed to cholera elimination in line with GTFCC guidance [1]. Because of the risk of re-introduction of cholera following elimination at ward and district levels [29,30], interventions are best planned using a decentralized community-centric approach to surveillance and case management through case-area targeted interventions. This has been seen to be effective in countries such as Uganda, DRC and Burundi, which are also all working towards elimination [7,61,62]. The predictable geographic location and seasonality of the outbreaks could be used to envisage the location and size of repeat vaccination campaigns with the possibility of pre-emptive campaigns timed before the rainy season to be included in the expanded program for immunizations [63–65]. Excitement surrounds the recent WHO prequalification of Euvichol S®, a simplified version of the Euvichol Plus® that is easier to produce but just as efficacious as its predecessors. [66] It is anticipated that its introduction in the global stockpile will increase vaccine access and hence permit countries, such as Zambia to plan for multiyear vaccination campaigns, as part of the cholera control and elimination efforts. These multiyear vaccine campaigns would serve as a bridge to increased WASH investments.

Similarly, these WASH investments can be planned in a decentralized construct as the different localities, even within the same country, have peculiar challenges expected to increase with the changing weather patterns [9,14,20,17,25]. Likewise, case management would need to have a further investigation into patient-specific factors such as host genomics. Research into the host microbiome is in early phases with mixed results but gives potential for newer treatment modalities such as probiotics and phage therapy against Vibrio cholerae [67-69]. Efforts to combat vaccine hesitancy, myths and misconceptions, must be made continuously, not only at the onset of outbreaks. This would help counter fears from the communities, particularly concerns of Western malevolence [45,46]. Here, we propose the need for enhanced collaborations of medical education and global exchanges of knowledge from low-income to high-income countries which would allow for hastened vaccine manufacturing locally, as recommended by the President of the Republic of Zambia, in his capacity as the World Health Organisation (WHO) global and Southern Africa Development Community (SADC) regional cholera control champion (70).

Isolates from the 2023/2024 outbreak in Zambia have yet to be fully analysed for host and pathogen genomics. However, recent sequencing results from Malawi give insight into a possible new transmission event into the subcontinent, that bears close resemblance to strains of Asian origin [71]. Zambia and Malawi share many porous borders, and trade and intermarriage are common between the people. The Malawian study postulated that the strain of vibrio in their 2022/2023 outbreak, the worst in Malawian history, was a highly successful cone of pandemic potential worsened by humanitarian and climate crises and then propagated by suitable environmental factors [71]. This agrees with earlier findings suggesting that outbreaks in Kanyama and other hotspots like the fishing villages, were due to a combination of recent introduction of newer pathogenic strains, and favourable environmental factors like deplorable WASH status [29, 30]. This also underscores the importance of joint cross-border surveillance and response activities in the region [21,30,31,32].

Challenges and gaps persist in cholera elimination efforts in Zambia. The need for a multisectoral, decentralised approach is evident, as no single intervention would remove all the various identified risk factors. The studies reviewed showcased different aspects of interventions during outbreak settings, or vaccination efforts in a reactive response. What can be seen is that cholera outbreaks in Zambia are progressively larger [8,20,23] and call for enhanced multisectoral and cross border collaboration [8,25]. Without environmental source control such as improving flush-to-sewage plumbing systems and overall climate-resilient solutions, it can be anticipated that the number of outbreaks in the country will continue to increase [11,14, 72, 73]. Our findings broadly confirm the need to align health and WASH investments with the GTFCC’s Roadmap to Cholera Elimination by 2030 [1] but also highlight the need for additional research across the various pillars to ensure tailored solutions, are adaptable to the local setting and able to inform best practice.

The study is limited mainly because of the scoping review methodology. Statistical tests could not be applied to any of the findings, and the overall recommendations are largely descriptive, differing from a systematic review. However, the challenges and gaps in cholera elimination in Zambia have been clearly outlined, and targeted interventions can be planned to ensure that the next iteration of the National Cholera Control Plan will be met with additional successes. Potential areas for future research on cholera elimination efforts must address best approaches for implementing community-centric surveillance and Case Area Targeted Interventions (CATI). Patient-level information on survival and transmissibility is lacking. There is a need to better understand the co-morbid conditions that may negatively influence patient outcomes, especially in patient groups such as the elderly or pregnant women. The role of host genetic factors such as the gut microbiome and its influence on symptomatology and transmissibility at the household level needs better understanding. Further, application of metagenomics to point-of-care testing to improve surveillance and how it relates to clinical outcomes remains an important gap in knowledge. The use of adjuvant therapies for vaccination and treatment is also not discussed in literature. Lastly, the impact of climate change on health in general and WASH interventions warrants a deeper dive to track the mechanisms and causal effects; be it extreme dryness due to drought or floods due to excess water. These are important gaps in knowledge for the global scientific community to consider as we work our way towards the 2030 elimination timeline.

## Conclusion

This scoping review collated evidence supporting a decentralised approach to cholera control in Zambia and Sub-Saharan Africa overall. Two key findings emerge from the analysis: first is the steady increase in cases and deaths over the years, despite adopting the first iteration of the Multisectoral Cholera Elimination Plan in 2019, and an anticipated increase in the coming years with rapid population growth and changing climate. The second key finding is that a wealth of evidence has already been generated in Zambia regarding best practices towards cholera control. There is a continued need to advocate strongly for multisectoral interventions with an alignment of health and WASH investment at the district and ward level, to align with this decentralised approach. The findings suggest many areas of further research considering the now endemicity of cholera in Zambia. We propose that our insights and recommendations can inform policymakers in crafting guidelines for implementing ward-level interventions, and these will be integrated into the next iteration of the National Cholera Control Plan. It is our hope that the lessons from here can be applied in other sub-Saharan African countries facing similar challenges and seeking to internalize the Global Roadmap for Cholera Control by 2030.

## Acknowledgements

This work is part of ongoing efforts from the Zambia National Public Health Institute, as the Secretariat of the National Cholera Control Taskforce, to better understand efforts towards Cholera Control in the Region. Many thanks to various task force members and partners who were directly and indirectly involved in this work.

## Data availability statement

All relevant data are included in the paper or its Supplementary Information.

## Author Contributions

### Conceptualization

Nyuma Mbewe, Nathan Kapata, John Tembo, Martin Peter Grobusch

### Methodology and Data Curation

Nyuma Mbewe, Nathan Kapata, William Ngosa, John Tembo, Martin Peter Grobusch

### Supervision and Validation

Nathan Kapata, Kelvin Mwangilwa, Mpanga Kasonde, Roma Chilengi, Kennedy Lishimpi, Lloyd Mulenga and Martin Peter Grobusch

### Writing, reviewing and editing

Nyuma Mbewe, John Tembo, Kelvin Mwangilwa, Paul Zulu, William Ngosa, Lloyd Mulenga, Kennedy Lishimpi, Joseph Adive Sereki, Mpanga Kasonde, Roma Chilengi, Nathan Kapata, and Martin Peter Grobusch

